# Saliva-based detection of COVID-19 infection in a real-world setting using reagent-free Raman spectroscopy and machine learning

**DOI:** 10.1101/2021.09.21.21262619

**Authors:** Katherine Ember, François Daoust, Myriam Mahfoud, Frédérick Dallaire, Esmat Zamani, Trang Tran, Arthur Plante, Mame-Kany Diop, Tien Nguyen, Amélie St-Georges-Robillard, Nassim Ksantini, Julie Lanthier, Antoine Filiatrault, Guillaume Sheehy, Caroline Quach, Dominique Trudel, Frédéric Leblond

**Author notes:** First author (Katherine Ember). These authors contributed equally to the work (François Daoust, Myriam Mahfoud).

## Abstract

**Significance:** The primary method of COVID-19 detection is reverse transcription polymerase chain reaction (RT-PCR) testing. PCR test sensitivity may decrease as more variants of concern arise.

**Aim:** We aimed to develop a reagent-free way to detect COVID-19 in a real-world setting with minimal constraints on sample acquisition.

**Approach:** We present a workflow for collecting, preparing and imaging dried saliva supernatant droplets using a non-invasive, label-free technique – Raman spectroscopy – to detect changes in the molecular profile of saliva associated with COVID-19 infection.

**Results:** Using machine learning and droplet segmentation, amongst all confounding factors, we discriminated between COVID-positive and negative individuals yielding receiver operating coefficient (ROC) curves with an area under curve (AUC) of 0.8 in both males (79% sensitivity, 75% specificity) and females (84% sensitivity, 64% specificity). Taking the sex of the saliva donor into account increased the AUC by 5%.

**Conclusion:** These findings may pave the way for new rapid Raman spectroscopic screening tools for COVID-19 and other infectious diseases.

## 1 Introduction

### 1.1 The current state of the COVID-19 pandemic

COVID-19 has precipitated the deaths of 3.9 million people worldwide as of the end of June 2021 and is the world’s most costly health crisis to date. The cost of the pandemic to the United States economy is projected to amount to $16 trillion.^1^ In a 19-month period, the SARS-CoV-2 virus has infected over 182 million people and numbers of confirmed cases continue to grow.^2^ Key difficulties in controlling the spread of SARS-CoV-2 are the long pre-symptomatic period, the wide range of non-specific symptoms, and the fact that asymptomatic individuals may be contagious.^3^ As governments worldwide have issued lockdowns and travel bans, rapid viral screening has become a crucial method to limit the spread of SARS-CoV-2.

### 1.2 RT-PCR as a gold standard

The current gold standard for SARS-CoV-2 testing is real-time reverse transcription polymerase chain reaction (RT-PCR) using oro-nasopharyngeal swabs. RT-PCR is a molecular biological diagnostic tool based on nucleic acid sequence detection. This uses nucleic acid primers, enzymes and cycles of heat to amplify a specific genomic sequence (from the SARS-CoV-2 genome in this case), enabling it to be detected more easily.^4^ Despite the fact that RT-PCR has successfully been used in testing for respiratory diseases, this method can show lower sensitivity for SARS-CoV-2 detection depending on the clinical presentation of the disease. Identifying asymptomatic patients early in the infection can help prevent and control the spread of COVID-19, so RT-PCR may fall short as a tool for mass serial screening of asymptomatic populations.^4,5^ Additionally, this method of testing requires complex tailored reagents which may be in short supply and lose specificity as the virus mutates. This has already been shown to be the case with influenza.^6^ To date, the Center for Disease Control has identified 13 variants of concern (VOCs) or variants of interest (VOIs) ^7^ and more will undoubtedly arise. We need to investigate alternative reagent-free approaches to reliably detect cases to control outbreaks and limit community spread of the disease.^8^

### 1.3 Saliva as a biosample

There are two primary ways of collecting SARS-CoV-2 ribonucleic acid (RNA) from a patient: nasopharyngeal (NPG) swabs and saliva sampling. Nasopharyngeal swabs are invasive and uncomfortable. There is also an increased risk for healthcare workers to be exposed to viruses due to patients sneezing or coughing.^9^ Meanwhile, saliva can be self-collected using non-invasive means, making it a much more attractive option for frequent screening. Saliva has been used successfully as a diagnostic tool for other coronavirus infections as the oral and nasal cavities can act as points of entry for respiratory viruses.^10^ SARS-CoV-2 may also enter the saliva from debris of the nasopharyngeal epithelium which drains into the oral cavity or from infection of the salivary glands.^11^ Consequently, using saliva samples paired with a reagent-free alternative approach could be an option for COVID-19 detection.

### 1.4 Raman spectroscopy

Raman spectroscopy is a light-based technique which measures the inelastic light scattered by matter, also called Raman scattering.^12^ This phenomenon was predicted in 1923 by Adolf Smekal and observed experimentally in 1928 by C.V. Raman and K.S. Krishnan^13,14^ but the potential biomedical application of Raman spectroscopy did not emerge until 1970.^15^ Raman scattering occurs when there is an exchange of energy between a sample and a monochromatic laser source emitting either visible or near-infrared light. The exchange of energy results in scattered light photons whose wavelengths are shifted compared to the excitation source in a way that depends on molecular structure and bonding. The difference in wavelength of the Raman scattered light compared to the incident light is called the Raman shift and two different effects can be observed: the Stokes shift (red-shift) or anti-Stokes shift (blue-shift).^12,16^ The Raman shift is usually measured in wavenumbers (cm-1) which are units that are inversely proportional to wavelength. Since the ground state is more populous at thermal equilibrium, the Stokes shift is more prevalent and commonly used in Raman spectroscopy.^16,17^

In a Raman spectrometer, a detector is used to measure light that has been inelastically scattered from a sample after it has passed through a series of filters, and these measurements are converted into a Raman spectrum. This is a plot of the intensity of the scattered light against the Raman shift in wavenumbers.^18^ Intensity of a Raman peak at a particular Raman shift increases as the concentration of molecules responsible for the peak increases. A Raman spectrum can therefore be thought of as a molecular fingerprint giving information about the molecules present in the sample through the analysis of the position, height, and width of peaks present in the spectrum. Raman spectroscopy can identify biomolecular features (such as lipids, proteins, nucleic acids, and amino acids) within biological samples. In recent years, clinical applications of this light-based technique have gained traction in oncology,^19–21^ inflammatory diseases,^22,23^ transplantation^24^ and virology.^25,26^ For example, Camacho et al.^25^ developed surface enhanced Raman sensors for the detection of the Zika virus by functionalizing core-shell nanoparticles with Zika ZIKV NS1 antibodies and reported a sensibility of 10 ng/mL.

### 1.5 Metabolic changes

While not all metabolic impacts of SARS-CoV-2 are known, some have been identified or can be anticipated from similar diseases. The severe acute respiratory syndrome (SARS) virus, SARS-CoV-1, has been shown to induce long-term metabolic changes in formerly infected patients. Elevated serum levels of cysteine, alanine, aspartic acid, succinic acid, and lactic acid were observed.^27^ The transmission vectors of the SARS-CoV-2 virus have been identified to be through the propagation of aerosols of biofluids such as saliva carrying a viral load.^28,29^ Reports have shown the high expression of angiotensin-converting enzyme II (ACE2) in epithelial cells of the oral cavity,^30^ ACE2 being a receptor which is involved in the mechanism of entry of the virus into cells. Furthermore, it has been hypothesized that COVID-19 may cause bacterial infection and other diseases of the salivary gland.^31^ This could theoretically be detected by Raman spectroscopy, as the concentration of analytes is correlated with the intensity, width, and specificity of Raman peaks.

### 1.6 Point-of-care rapid screening

In a pandemic context, fast near real-time screening is required to track and contain the propagation of a virus. Airports, schools, workplaces, and remote communities would benefit from a rapid, reagent-free screening technique. The sensitivity and the label-free approach of Raman spectroscopy makes it a candidate for fast, robust, low-cost, and transportable means of viral screening to complement the diagnosis capacity of the gold standards. Any COVID screening tool should be applicable to all users, regardless of symptoms, age, sex, time-of-day of sample collection or diet. This poses a challenge when developing a label-free Raman spectroscopic approach as saliva composition is affected by time of day, sex, age and potentially other underlying health conditions.^32– 38^

A 2021 study (n = 30 for COVID positive volunteers) found that Raman micro-spectroscopy could detect COVID-19 infection in saliva with 84% sensitivity and 92% specificity.^39^ However, the clinical cohort was restricted to elderly volunteers who had presented to hospitals and the study design required saliva collection to occur prior to breakfast. Thirty-six percent of positive cases were severely or critically ill, and a further 30% had symptoms and evidence of pneumonia upon imaging. Another recent study (n = 29 for COVID positive volunteers) determined infrared spectroscopy could be used to discriminate between COVID-infected and non-infected saliva with 93% sensitivity and 82% specificity.^40^ As for the previous study, both the COVID-positive and negative cohorts consisted of hospitalized, symptomatic patients requiring treatment. Most label-free tests would be aimed at screening non-hospitalized people who may be symptomatic or asymptomatic with different levels of severity of COVID-19 infection. Sample collection information and other clinical characteristics apart from viral load were not reported for this dataset, including age, sex and comorbidities. To be applicable in a setting outside of hospitals, a label-free screening technique must be applicable to people of all ages, with or without symptoms, regardless of diets, smoking status, with samples collected at any time of day.

Here, we present the results of a study demonstrating that Raman spectroscopy combined with machine learning is a candidate for real-world COVID-19 screening in the general population. The study design was developed to ensure that the number of samples collected allowed us to match COVID-positive and COVID-negative samples in terms of potential confounding factors including sex at birth, age, COVID symptoms, body mass index and prescription drugs taken. We studied saliva samples taken from volunteers at a COVID-19 testing clinic, including asymptomatic volunteers and those with respiratory and non-respiratory symptoms. Samples were taken from people aged >10 years old to <61 years old.

## 2. Experimental

### 2.1 Sample collection

The experimental workflow is shown in Figure 1. A total of 37 COVID-19 positive and 513 COVID-19 negative samples were collected from the Pointe-Saint-Charles COVID-19 testing clinic in accordance with ethical guidelines from the Centre Hospitalier de l’Université de Montréal (CHUM) Research Ethics Board (project number: 20.133). Volunteers presenting between 10 am and 2 pm were asked to complete a questionnaire reporting their symptoms and associated biological and environmental factors that could affect saliva composition e.g., age, sex at birth and comorbidities. When applicable, volunteers were asked to remove any lipstick or lip balm using makeup removal wipes (About Face Cleansing Wipes, Micronova Manufacturing Inc., Torrance, California). Each volunteer was instructed to first rinse their mouth three times with bottled water to remove food debris. Volunteers then waited 5-10 minutes for saliva to accumulate before spitting in a 50 ml Falcon tube to collect a minimum of 1.5 ml of the biofluid. Tubes were then immediately stored in a refrigerator at 4°C. Samples were transported to the Centre de Recherche du CHUM (CRCHUM) on ice. Samples were classified as being from asymptomatic, non-respiratory symptomatic or symptomatic volunteers based on symptoms reported in the questionnaire. “Respiratory” symptoms included those that might result in significant amounts of mucus in saliva such as runny nose, difficulty breathing, sore throat, coughing or wheezing. “Non-respiratory” symptoms included all other symptoms including headache, muscle aches and tiredness. The classification of each sample as being obtained from a COVID-19 positive or negative volunteer was based on PCR tests either based on nasopharyngeal or saliva swabs.

**Figure 1:**
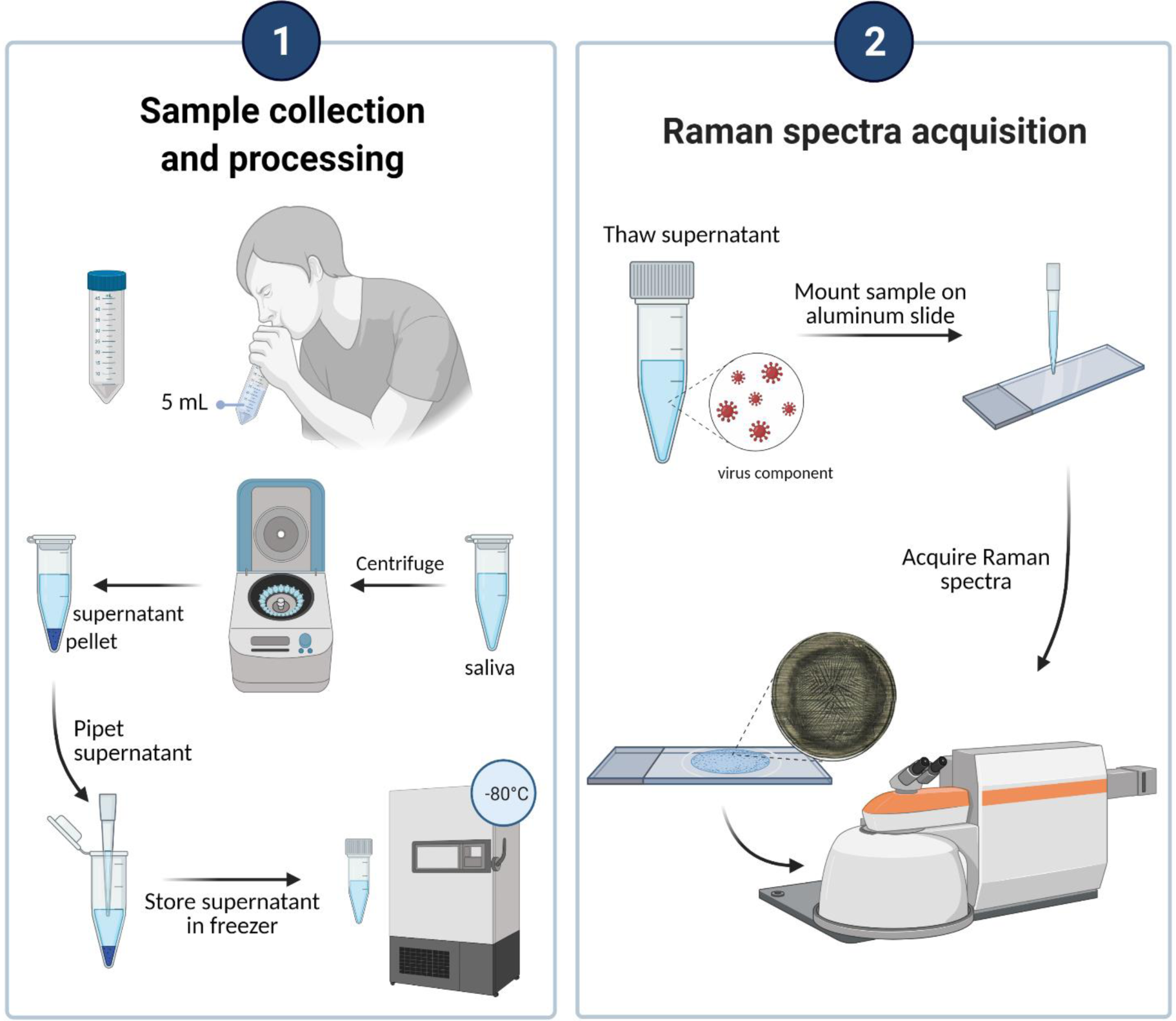
Workflow from saliva collection to determination of COVID infection status from Raman spectra. Volunteers donated between 1 and 5 ml of saliva into a 50 ml tube. The liquid was pipetted into a 1.5 ml microcentrifuge tube which was centrifuged. The saliva supernatant was then stored at -80°C. Supernatant was thawed, vortexed and mounted on an aluminum slide. After 45 minutes of drying, spectra were acquired using a Renishaw InVia Raman spectrometer. This figure was created with BioRender.com.

### 2.2 Pre-imaging sample processing protocol

Saliva samples were processed and imaged at biosafety containment level 2 (BSL2) in biosafety cabinets. 1 ml of whole saliva samples were transferred to 1 ml microcentrifuge tubes and centrifuged at 4000 rpm for 30 minutes at 4°C. The supernatant was pipetted into one cryotube, mixed and then 40-500 µl were aliquoted into five separate 1.8 ml cryotubes. The part of the supernatant near the pellet was discarded and the pellet was retained in the microcentrifuge tube. Pellet and supernatant samples were stored at -80°C. Prior to Raman spectroscopy interrogation, saliva supernatant samples were thawed at room temperature for 30 minutes, vortexed for 40 seconds and 10 µl were pipetted onto an aluminum slide. Droplets were allowed to dry at room temperature for at least 45 minutes.

### 2.3 Model saliva preparation

Model saliva was made using concentrations listed in Supplementary Table S1. As for human saliva supernatant, it was frozen at -80°C and thawed at room temperature for 30 min, vortexed for 40 s and 10 µl of sample were pipetted onto an aluminum slide and dried. Bovine submaxillary mucin was used to generate the model saliva due to limitations in availability of human mucin. To address this limitation, Raman spectra from a limited available quantity of human mucin type I were also acquired for more accurate human mucin peak assignment in human saliva supernatant samples.

### 2.4 Raman microspectroscopy of dried saliva samples

Dried saliva supernatant samples and model saliva were imaged using an inVia™ confocal Raman microscope (Renishaw, Gloucester, UK) in reflection mode. Brightfield montage images of each droplet were obtained with a 5x lens. Brightfield montage images of the central crystalline region and edge region were obtained using a 50x objective. For Raman spectral acquisitions, the excitation consisted of a 785 nm 40 mW laser in line-focus mode (3 µm x 8 µm spot size) with a 1200 l/mm grating. Spectra were acquired in the fingerprint region (between 602 and 1726 cm^-1^) from three separate morphological regions within each droplet: “edge” (the perimeter of the droplet), “on crystal” (inside visible crystals in the center of the droplet) and “off crystal” (outside visible crystals in the center of the droplet). Ten spectra were acquired in each region and 8-10 repeat measurements were taken from each point. Spectra were taken from random “on crystal” and “off crystal” regions in the densest region of the droplet, and in a zig-zag pattern within the edge to capture spectra from the very edge and slightly closer to the center (Figure 2). Each measurement was made ensuring 60-70% of the Raman microscope sensor dynamical range was used to minimize the impact of shot noise, resulting in an acquisition time of 2-10 s for “edge” and 15-40 s for center spectra (“on crystal” and “off crystal”), depending on the level of sample autofluorescence.

**Figure 2:**
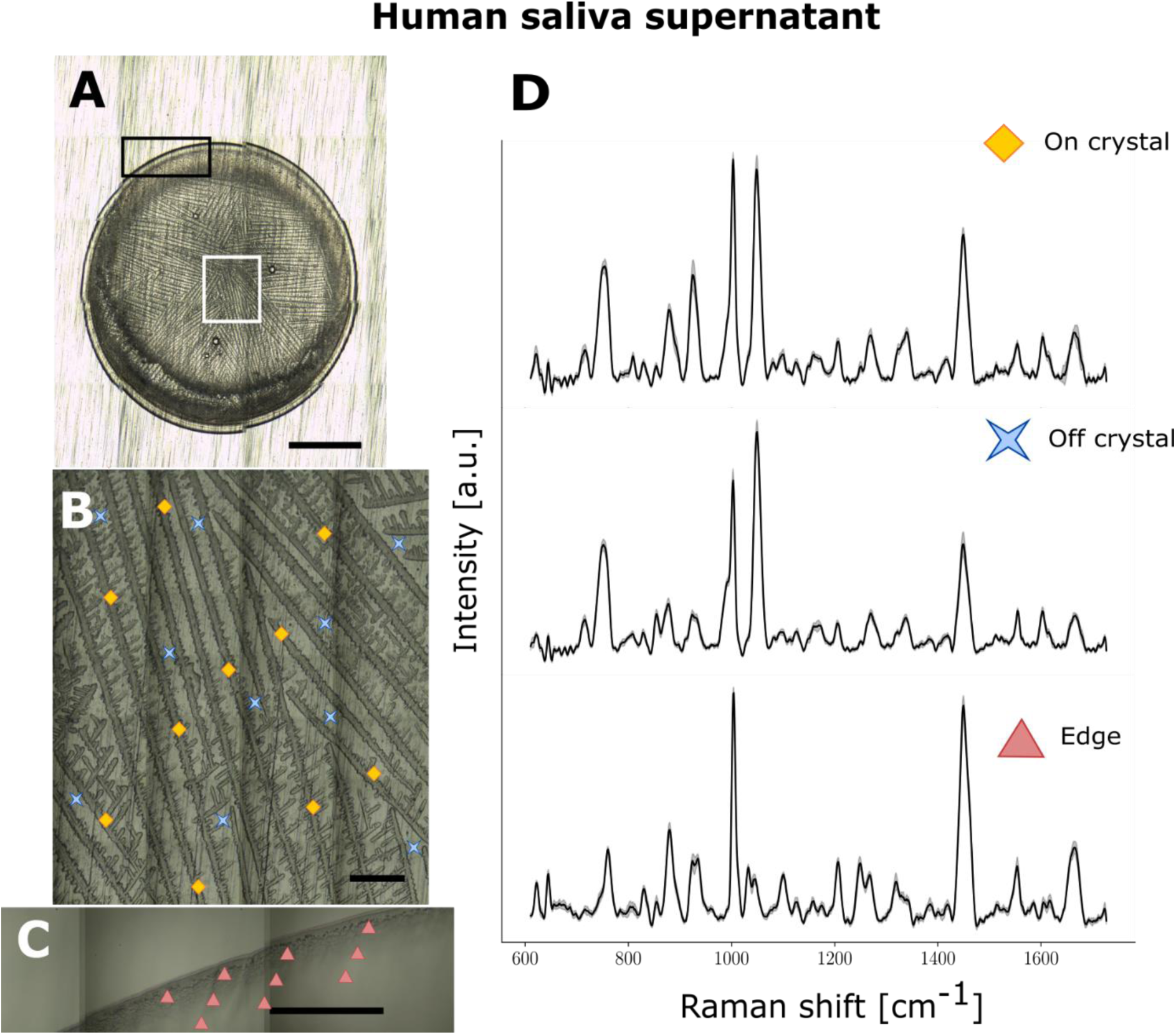
Raman spectroscopy of a representative droplet of human saliva supernatant from a COVID positive volunteer. (A-C) Brightfield images for (A) whole droplet dried on aluminum slide (5x), (B) crystalline region (50x) and (C) edge (50x) with acquisition points shown with different symbols: diamonds (“on crystal”), crosses (“off crystal”) and triangles (edge). (D) Average spectra from one saliva sample for “on crystal” (top), “off crystal” (middle), and “edge” (bottom) regions respectively with shaded areas representing the interspectral variance within the specimen. Each spectrum is an average of multiple acquisitions obtained with a 785 nm laser using a Renishaw InVia Raman microscope. The scale bar in A is 1 mm in length, whilst the scale bar in B and C are 0.1 mm long.

### 2.5 Raman microspectroscopy of individual model saliva components

Solid compounds placed on aluminum slides were imaged using the Renishaw InVia microscope. 25 spectra were taken from each compound using the 50x lens between 105-1727 and 2601-3359 cm^-1^.

### 2.6 Spectral data processing

The following data preprocessing steps were applied to every individual measurement: 1) removal of cosmic rays was imperative due to the long acquisition time, 2) smoothing using a Savitzky-Golay filter of order 3 with a window size of 11, 3) background subtraction of signals produced by the aluminum slides and autofluorescence using a custom adaptation of the rolling ball algorithm ^41^, 4) cropping the region below 1100 cm^-1^ due to the large variances at lower wavenumber shifts, 5) averaging of the repeat measurements taken at a given spatial point, and 6) standard normal variate (SNV) normalization.^21^

### 2.7 Data preparation and feature selection

For data quality reasons, a limited number of spectra (from 3 patients for “on crystal”, from 5 patients for “edge”) were removed before training the machine learning models. Exclusion was based on abnormally high levels of residual stochastic noise (after background removal) or the presence of unusual spectral shapes unrelated to biomolecular content. For each spectrum, a Gaussian fitting procedure was applied to all peaks of biological origin fitted to extract its position, its height, and its width; a total of 24 peaks were extracted using this procedure. These –along with the relative intensity of 700 individual bands in a spectrum– represent the data from which machine learning models could be trained.

A feature selection technique was applied to reduce the size of the feature set prior to model machine learning model training. Feature reduction is necessary because some spectral regions either provide no useful information for the classification or are perhaps too correlated. Algorithms typically run with at least one parameter, known as hyperparameter, that needs to be initialized. A variance-based algorithm was used to coarsely reduce the number of features. Features presenting a large variance have a higher weight and by sorting them according to their weight, only the k-best features are retained, where k was varied between 5 and 80 for the spectral features and 5 and 72 for the peak features. A Random Forest classifier (RF) with 200 estimators is then used to further reduce the feature set, this time with a multivariate technique rather than a univariate one. Again, this was done independently for spectral and peak features. In the end, the final number features will range between 20 and 200, with an average of 90.

### 2.8 Classification model and statistical analyses

A multiple-instance learning (MIL) approach was favored to be at the core of the classification algorithm. In such a scenario, all the spectra (instances) belonging to a given patient are represented by a bag to which a label is associated (COVID positive or negative) rather than labelling the instances individually. Labels associated to each patient (bag) correspond to the result from their PCR test. More specifically, the selected method is a multiple-instance learning via embedded instance selection (MILES) algorithm that compute a similarity measure between instances to map them into a different feature space ^42^. Instead of being represented by multiple instances from the original feature space, each bag is now represented by a single instance, based on the similarity measure from the new feature space. Under MILES, the mapping is done using an *intermediate instance pool* (IIP) which contains all the instances. It is also possible to create a *discriminative instance pool* (DIP) which is a subset of the IIP represented only by the best instances. This method is called a MIL with discriminative mapping (MILDM) algorithm ^43^. The similarity function has a hyperparameter, σ^2^, that varies between 1 and 70. For high σ^2^ values, the similarity function tends to 1, and 0 for small σ^2^ values. The size of the DIP is defined by a hyperparameter, m_dip_, that varies between 5 and 60% of the total number of spectra. Following the mapping, for both MILES and MILDM algorithms, the same feature selection algorithm used for peak and peak features is applied to the new mapped feature space. The number of features in the MILES and MILDM spaces are defined by hyperparameters k_MILES_ and k_MILDM_ that vary between 5 and 60% of the total number of spectra, and between 5 and m_dip_, respectively. A RF classifier with 200 estimators is applied subsequently. Once the bags have been mapped to the new feature space, either with MILES or MILDM, a standard Support Vector Machine (SVM) algorithm is used for the training. Because we used a linear SVM, the only hyperparameter is the regularization parameter *C* which corresponds to the penalty term and was varied between 1 and 50.

As the values for the hyperparameters of the feature selection and classification algorithms are not known *a priori*, many combinations need to be considered. A combination is formed by randomly selecting a value for each hyperparameter within their respective range. A 5-fold cross validation (CV) procedure is used to assess the classification performance for each hyperparameter combination. Once applied on the validation set, the trained model will output a classification probability, continuous between 0 and 1, for each sample in the validation set. This procedure is repeated until all folds have been used once as a validation set. The model performance is assessed with a receiver operating characteristic (ROC) curve from which we can compute the accuracy, sensitivity, and specificity associated to the point with the minimal distance to the upper left corner and the area-under-curve (AUC). The ROC curve is computed by comparing the classification probabilities from the validation sets to their pathological labels, either 0 (COVID negative) or 1 (COVID positive), by varying the threshold value between 0 and 1.

In order to test the repeatability of the results, all these steps were repeated five times, allowing the data to be split differently between the folds. The final performance assessment is an average ROC curve with its own AUC associated. This procedure is repeated until all the desired hyperparameter combinations have been considered and the set of hyperparameters with the highest performance corresponds to the final model.

## 3 Results

### 3.1 Saliva collection and preparation protocol, volunteer demographics

We developed a technique for obtaining Raman spectra from saliva supernatant in a way that minimized person-to-person variation (Figure 1). Raman spectroscopy is sensitive to all Raman-active molecules within a sample and so minimizing exogenous particles is critical. We had previously found that the Raman spectrum of chromophores was present in volunteers who donated saliva whilst wearing lipstick (Supplementary Figure S1). We therefore provided volunteers with lipstick removal wipes if necessary and implemented a mouth washing step to remove food debris. After a waiting time of 5 minutes, whole saliva was collected from 550 volunteers at a COVID-19 testing site and processed in a containment level 2 facility (clinical characteristics listed in Table 1). Saliva was centrifuged to remove further food particles and the supernatant stored at -80°C. The supernatant was allowed to thaw at room temperature. It was then vortexed, dried on an aluminum slide, and analyzed using a Raman microspectrometer (InVia, Renishaw, UK). Spectra obtained from aqueous phase saliva supernatant were primarily fluorescence with no visible Raman peaks, but upon drying we could obtain Raman spectra with clearly distinguishable Raman peaks. The methodology is described in the materials and methods section in more detail. Clinical characteristics of the volunteers are reported in Table 1. The viral loads of our samples ranged from a cycle threshold of 15.5 (very high) to 36.3 (very low) (Supplementary Table S1).

**Table 1:**
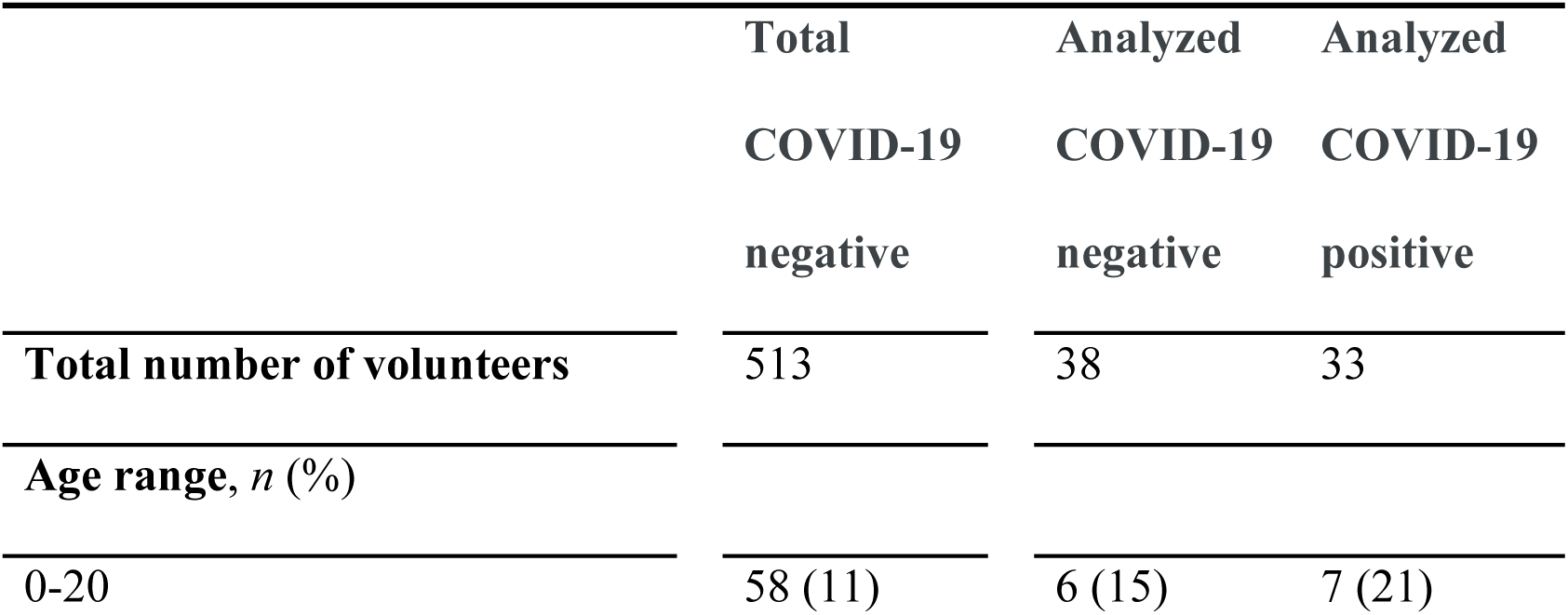
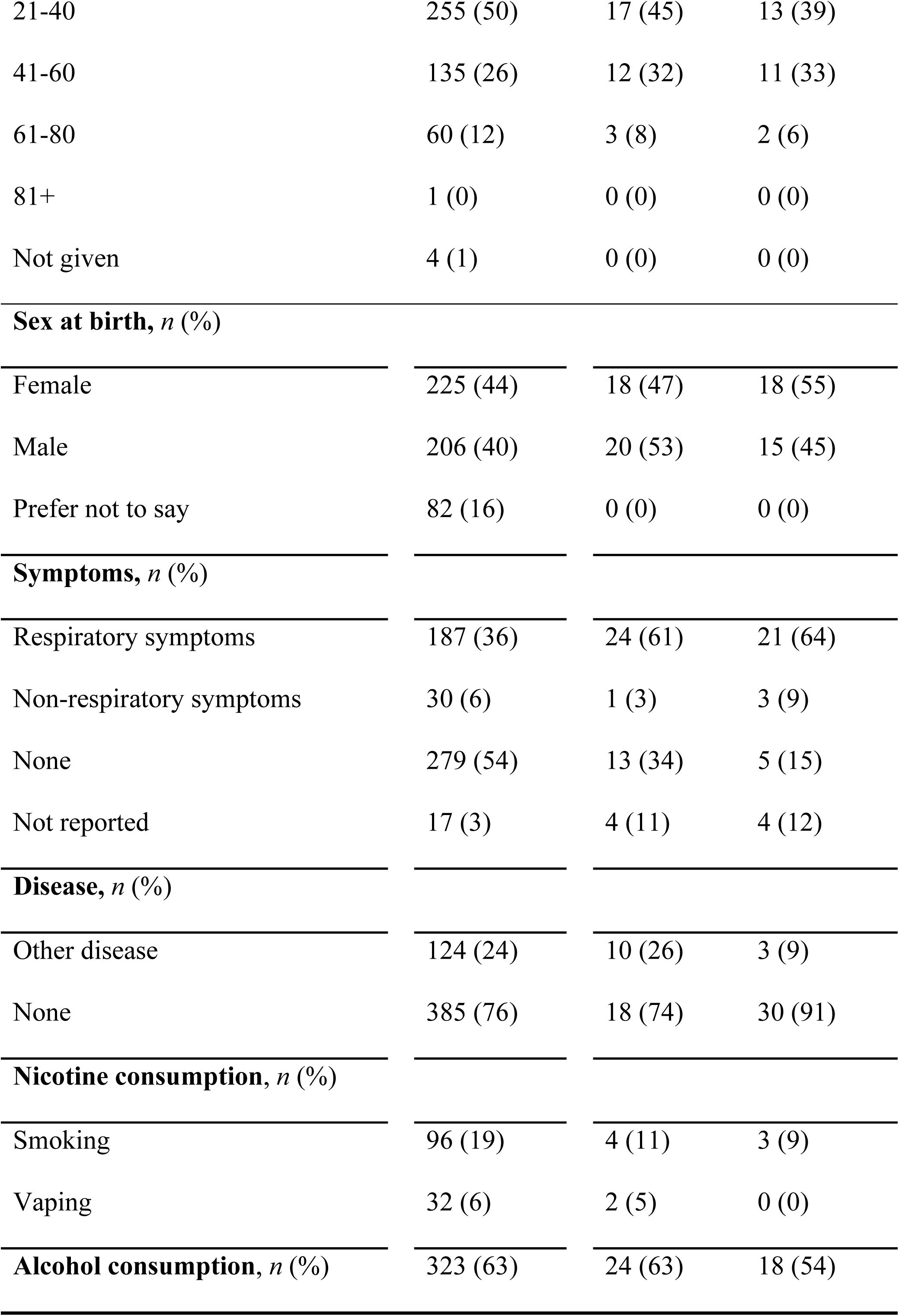
Clinical characteristics of the total volunteer cohort. Characteristics were taken from questionnaires given to volunteers. There were 513 COVID negative volunteers and 37 COVID positive volunteers. The number on the left in each column is the number of individuals with each characteristic, whilst the number in parentheses on the right is the percentage of the total number of COVID negative or positive volunteers.

### 3.2 Human saliva supernatant forms morphological regions with distinct Raman spectra

Raman microspectrometers are spectrometers coupled to microscopes to yield Raman spectra with a spatial resolution that can be modulated by the use of objectives with different magnifications. A single Raman spectrum can be obtained from a discrete point within a sample and the area of excitation in these experiments was 3 µm by 8 µm, using a 50x objective. In brightfield images (5x and 50x) taken with the system, we observed that drops of human saliva supernatant most frequently dried with a translucent crystalline region in the center and a slightly more opaque peak around the edge (Figure 2A-C). This is a common phenomenon observed in dried water-based droplets due to the “coffee-ring effect” in which suspended particles accumulate at the edge of the droplet. In some saliva droplets, there was also a region between the crystalline center and edge where there were no crystals and Raman signal was minimal which could be attributable to a lower concentration of Raman-active molecules.

In order to determine which part of the droplet should be interrogated for discrimination between COVID-negative and COVID-positive samples, we took measurements from both the center (Figure 2B) and the edge (Figure 2C) of the dried droplet. The central region was divided into “on crystal” – where measurements were taken from visible crystals – and “off crystal” – where measurements were taken from the milieu in-between crystals. Spectra were taken at 10 points in each of the “edge”, “on crystal” and “off crystal” regions with 8-10 successive acquisitions taken from each point to increase signal-to-noise ratio (SNR) by averaging the spectra (Figure 2D). In the “on crystal” region of human saliva, we observed strong peaks at 1003 and 1045 cm^-1^ as well as 853 and 930 cm^-1^ in some volunteers. In the edge region, we observed strong peaks at 1003, 1449 and 1655 cm^-1^ (Supplementary Table S3). These different Raman signatures indicate that the molecular content of the center and the edge of dried saliva supernatant is different. As a droplet dries, certain molecules accumulate at the edge and others in the center. The strong peak at 1003 cm^-1^ is common in Raman studies of biological materials as it corresponds to the ring breathing mode of phenylalanine and is often one of the strongest peak in proteins ^44^.

### 3.3 Raman peak assignment by comparison of human saliva to model saliva

To confidently assign Raman peaks to donor spectra, we used a saliva model adapted from Sarkar *et al*. ^45^ containing salts, bovine serum albumin, mucin and other metabolites. The precise composition of the model is listed in Supplementary Table S2. We dried the droplet in the same way as the human saliva supernatant samples and observed similar morphology in brightfield images (Figure 3A-C) as in human saliva. Although there are some particles of solid compounds which had not completely dissolved in the model saliva, the branched crystalline region resembles that of human saliva supernatant. The edge region of model saliva also resembles that of human supernatant in terms of size and the fact that it lacks crystals.

**Figure 3:**
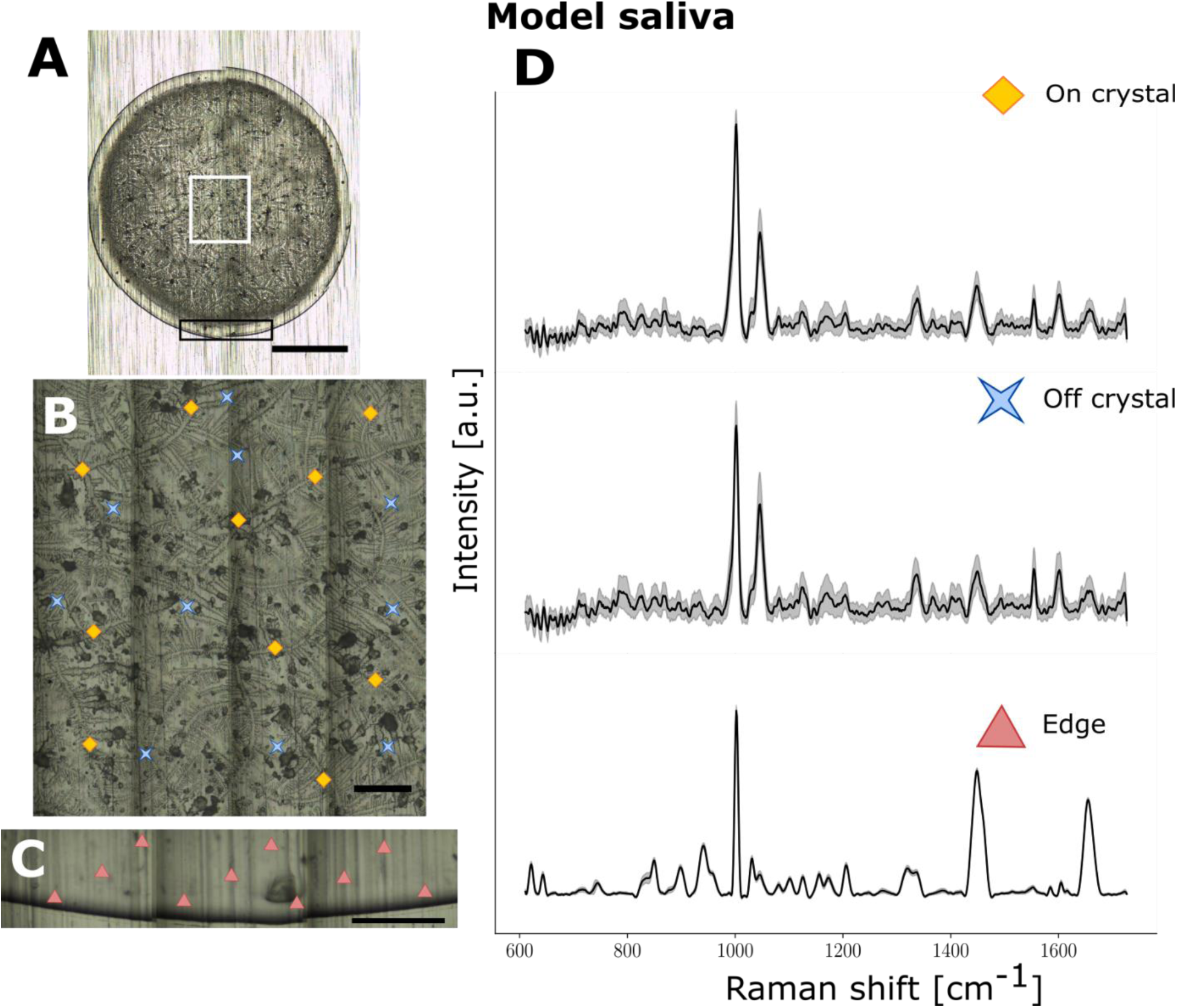
Raman spectroscopy from a droplet of model saliva composed of a mix of salts, bovine serum albumin, mucin and other metabolites. (A-C) Brightfield images for (A) whole droplet dried on aluminum slide (5x), (B) crystalline region (50x) and (C) edge (50x) with acquisition points shown with different symbols: diamonds (“on crystal”), crosses (“off crystal”) and triangles (edge). (D) Average spectra from one saliva sample for “on crystal” (top), “off crystal” (middle), and “edge” (bottom) regions respectively with shaded areas representing the interspectral variance within the specimen. Each spectrum is an average of multiple acquisitions obtained with a 785 nm laser using a Renishaw InVia Raman microscope. The scale bar in A is 1 mm in length, whilst the scale bar in B and C are 0.1 mm long.

Furthermore, the Raman peaks from the center and the edge of human saliva supernatant (Figure 3D) resembled those of model saliva, suggesting a similar molecular composition. We took spectra of the pure constituents of the model saliva to determine which molecules give rise to peaks in the model saliva and whether these peaks were also present in human saliva supernatant (Supplementary Figures S3-S6). By comparing spectra of human saliva supernatant to model saliva, we could identify Raman peaks due to phosphate, nitrate, proteins and nucleic acids (Supplementary Table S3). We found that peaks due to salts (nitrate and phosphate) were strong in spectra taken from the central crystalline region of both model saliva and real human saliva supernatant. Meanwhile, the spectra from the edge of the drop were primarily composed of peaks due to proteins (phenylalanine, amide II, amide III). This is consistent with findings in serum and other biofluids – proteins move towards the edge of the droplet in the drying process whilst salts may be spread throughout the droplet ^46,47^.

### 3.4 Predictive modeling for segmented saliva samples

To fully explore the potential of all regions of the dried saliva supernatant matrix for detection of COVID-19, we developed predictive models based on “edge”, “on crystal” and “off crystal” regions. The large number of COVID-negative saliva samples allowed us to match each COVID-positive sample with a negative sample having approximately the same characteristics in terms of sex at birth, age, COVID symptoms, body mass index (BMI) and prescription drugs taken (Table 1). This ensured that the clinical characteristics of the samples analyzed were approximately the same between the 33 positive (15 males, 18 females) and 38 negative (20 males, 18 females) samples. Patient matching is done to reduce the impact of potential confounding factors. However, there was a slightly higher percentage of analyzed samples from volunteers with comorbidities in the COVID negative group. Comorbidities included optic neuritis, cancer, hypertension, diabetes, anxiety, allergies and migraines.

Machine learning (ML) algorithms were trained using spectra from 33 COVID positive and 38 COVID negative samples. The area of laser excitation was small (24 µm^2^) relative to the area of the droplet (approximately 12 mm^2^). Due to the crystallization process and dilution of biomolecules within saliva, not all spectra necessarily carry molecular information relevant to the COVID status of the patient. As a result, a ML approach needed to be adapted whereby each spectrum acquired from a saliva droplet from a volunteer could be treated independently (Figure 4). The ML approaches that were employed are based on multiple instance learning (MIL); specifically, MIL via embedded instance selection (MILES) and MIL with discriminative bag mapping (MILDM). The implementation details of techniques are presented in the Experimental section. For each classification scenario presented below, results are shown using a receiver operating characteristic (ROC) curve for both MILES and MILDM.

**Figure 4:**
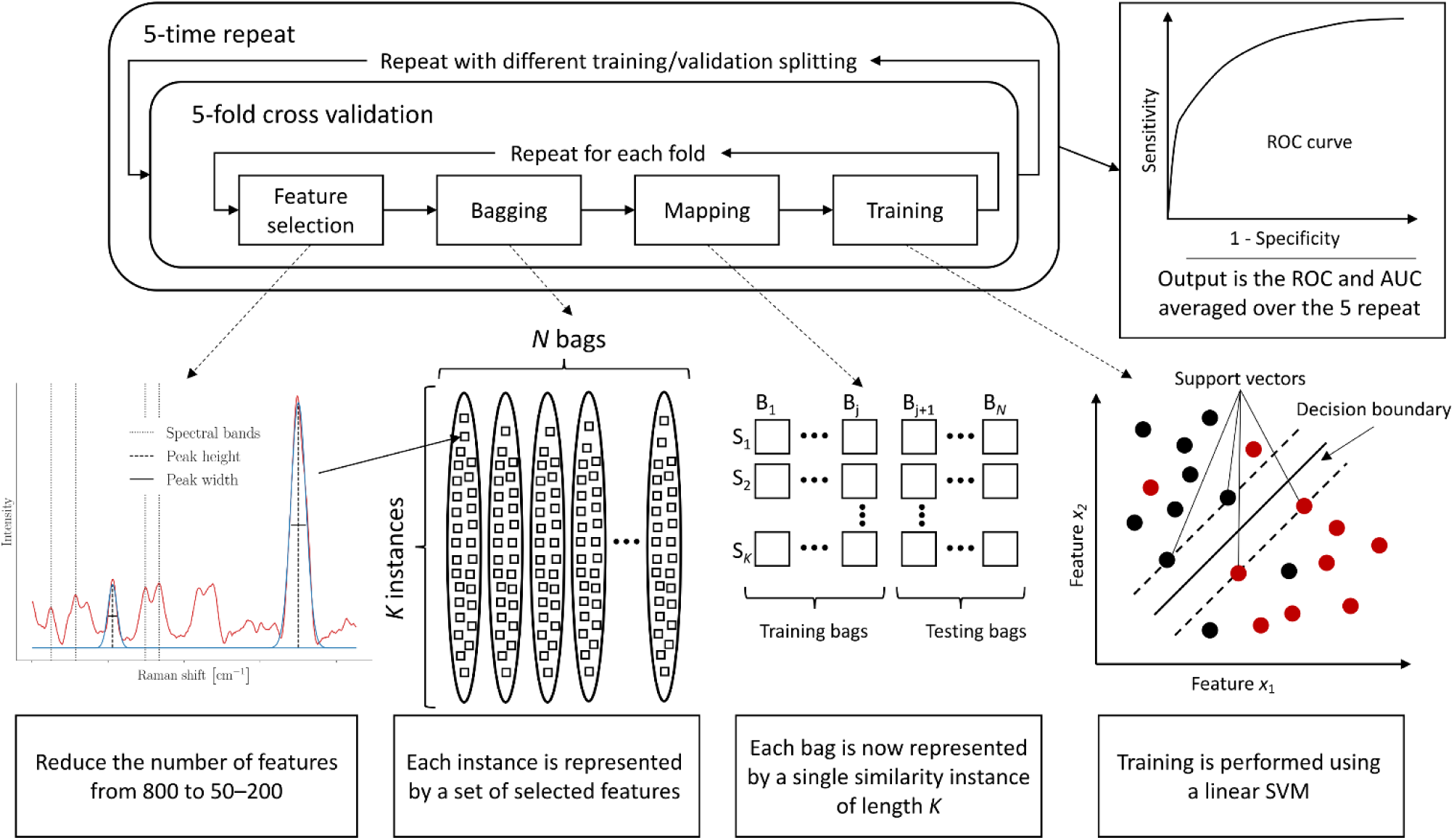
Machine learning schematic workflow. The ML workflow consists of a 5-fold cross-validation (CV) embedded in a 5-time repeat loop creating different splitting of the training and validation sets. Feature selection, bagging, mapping, and training steps are repeated for each fold. Raman spectra are represented by spectral peaks and fitted peaks. Each instance is represented by the relevant features which are selected using a combination of a variance-based algorithm, acting as a broad skimmer, and a random forest (RF). Each bag is then mapped from a multiple instance representation to a single instance representation through an instance similarity measure; the mapping function being different for MILES and MILDM. A linear Support vector machine (SVM) algorithm is used to train each model and output a classification probability for patients in the validation set. After each CV procedure, a receiver operating ROC curve is computed with a corresponding AUC value to assess the model performance. The final output is the ROC and AUC averaged over the 5 repetitions, ensuring further stability.

### 3.5 Separating samples based on sex at birth increases accuracy of COVID detection

As sex hormones can affect metabolism and immune responses and thus the molecular content of saliva, we separated samples based on sex at birth. In males, we could discriminate between COVID positive and negative saliva supernatant samples with a receiver operating characteristic (ROC) curve AUC of 0.80 using machine learning on Raman spectra taken from the “edge” region (Figure 5A&B). This predictive model was built using MILDM (*n* = 35, 15 COVID positive and 20 COVID negative samples), yielding a sensitivity of 79% and a specificity of 75%. Key features used in model building included peaks that can be assigned to carbohydrates, carotenoids, proteins and nucleic acids (Figure 5C). In males, models built using the “on crystal” region had a lower AUC than the edge models (0.72 with MILDM, 15 COVID positive and 20 COVID negative samples) (Figure 5D-F).

**Figure 5:**
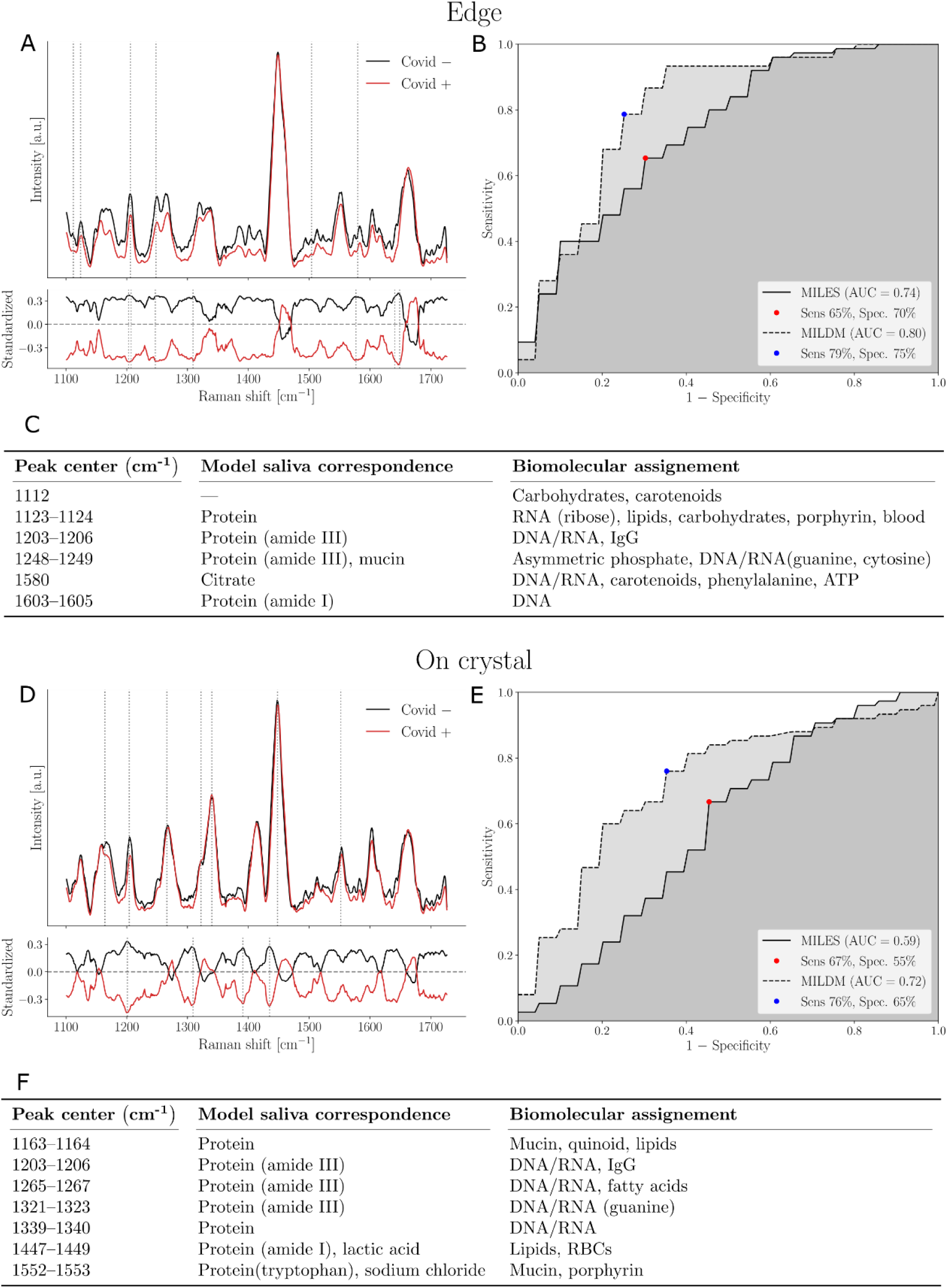
Machine learning model discriminating between COVID-negative and positive saliva supernatant from males using (A-C) “edge” and (D-F) “on crystal” Raman spectra from dried droplets. (A, D) Upper frame shows SNV-normalized, baseline corrected Raman spectra from all volunteers. Variance is shown by pale lines (variance of mean spectrum from each individual) and main features used in model building designated by dotted lines. Mean COVID-negative spectra (n = 20, at least 8 spectra per volunteer) are shown in black and COVID-positive spectra (n = 15, at least 8 spectra per volunteer) are shown in red. Bottom frame shows the standardized Raman spectra, where each individual feature has 0 mean and unit variance. (B, E) Receiver operating curve (ROC) for these models with sensitivity and specificity. (C, F) List of features used in model building and their assignments as determined using compounds in model saliva and from literature.

For females, using “on crystal” spectra, we could discriminate between COVID positive and negative saliva samples with an AUC of 0.80 (Figure 6D&E). This model was built using MILES (*n* = 36, 18 COVID-positive and 18 COVID-negative), with a sensitivity of 84% and a specificity of 65%. Key features included peaks that can be assigned to lipids, proteins and nucleic acids (Figure 6F). In females, models built using the edge region had a lower AUC than the “on crystal” models (0.67 with MILES, *n* = 36, 16 COVID-positive, 18 COVID-negative) (Figure 6A-C). This is reflected by the observation that the mean edge spectra of COVID-positive and COVID-negative females have much greater overlap than the mean edge spectra from males, suggesting the molecular composition of the protein-rich edge region of saliva is not altered as severely in females as it is in males.

**Figure 6:**
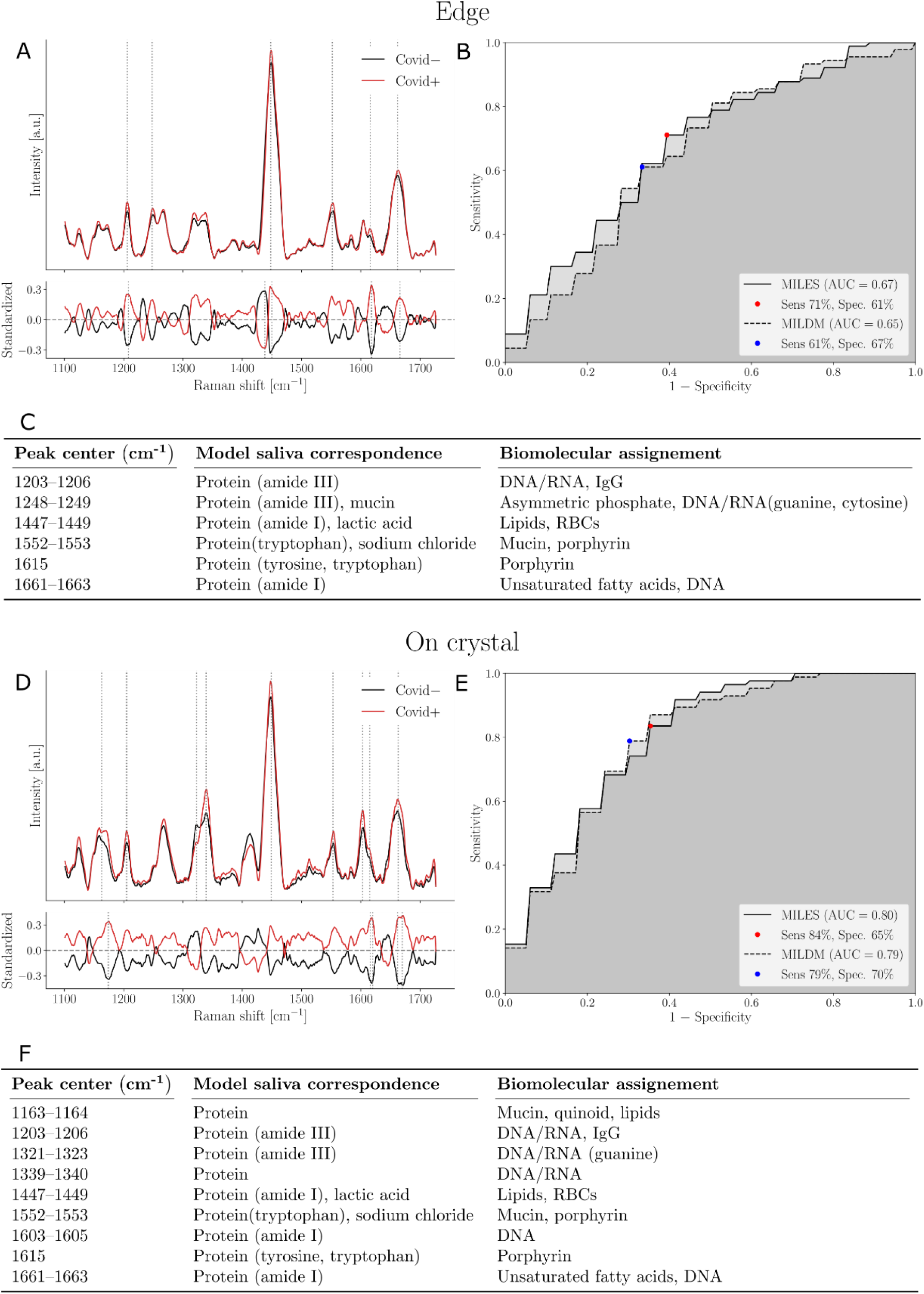
Machine learning model discriminating between COVID-negative and positive saliva supernatant from females using (A-C) “edge” and (D-F) “on crystal” Raman spectra from dried droplets. (A, D) Upper frame shows SNV-normalized, baseline corrected Raman spectra from all volunteers. Variance is shown by pale lines (variance of mean spectrum from each individual) and main features used in model building designated by dotted lines. Mean COVID-negative spectra (n = 18, at least 9 spectra per volunteer) are shown in black and COVID-positive spectra (n = 18 for edge, n = 16 for on crystal, at least 9 spectra per volunteer) are shown in red. Bottom frame shows the standardized Raman spectra, where each individual feature has 0 mean and unit variance. (B, E) Receiver operating curve (ROC) for these models with sensitivity and specificity. (C, F) List of features used in model building and their assignments as determined using compounds in model saliva and from literature.

The best model in males (built using the “edge”) only shared 1203-1206 cm^-1^ and 1603-1605 cm^-1^ with the best model in females only. All other features were different.

### 3.6 Discrimination between PCR-positive and PCR-negative samples regardless of sex at birth

Amongst all confounding factors including sex at birth, we could discriminate between COVID positive and negative saliva supernatant samples using machine learning on Raman spectra taken from the edge region with an AUC of 0.76 (Figure 7A&B). This predictive model was built using MILDM (*n* = 71, 33 COVID-positive and 38 COVID-negative), yielding a sensitivity of 73% and a specificity of 71%. This reduced the AUC by 0.04 relative to the model built in males alone using the same part of the droplet. Half of the top molecular features are shared between the two models (Figure 5C and 7C). We could discriminate between COVID-19 positive and negative “on crystal” regions with an AUC of 0.69 using MILES (*n* = 69, 31 COVID-positive and 38 COVID-negative) (Figure 7D&E). This resulted in a reduction in AUC of 0.11 compared to the model built in females only using the same part of the droplet (Figure 6E). More than 65% of the top features are shared between the two models (Figures 5C and 7C). We could not build a reliable model using spectra from the “off crystal” region (AUC = 0.57). This was true for all models built, so the “off crystal” region was removed from the analyzes.

**Figure 7:**
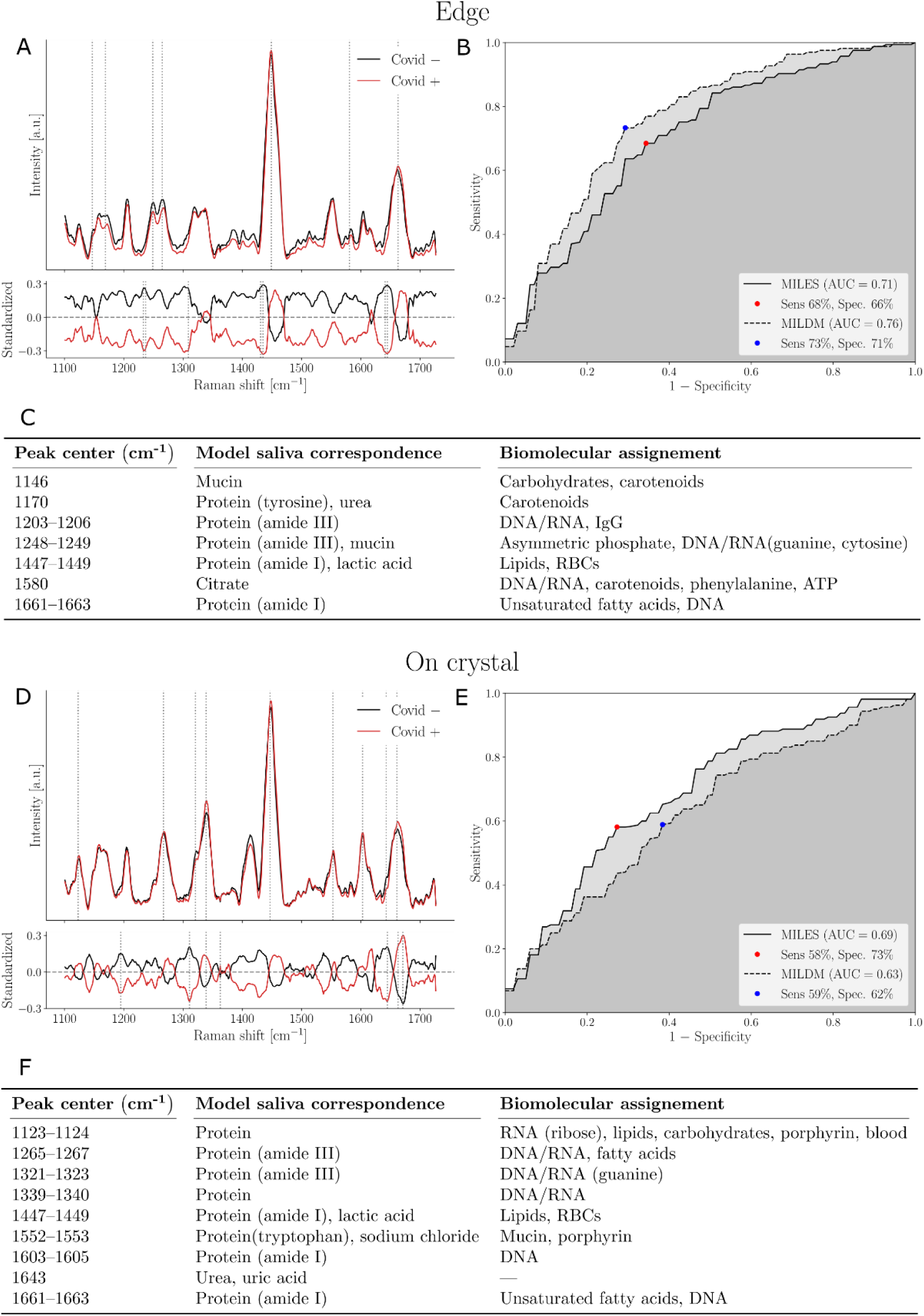
Machine learning model discriminating between COVID-negative and positive volunteer saliva supernatant using (A-C) “edge” and (D-F) “on crystal” Raman spectra from dried droplets. (A, D) Upper frame shows SNV-normalized, baseline corrected Raman spectra from all volunteers. Variance is shown by pale lines (variance of mean spectrum from each individual) and main features used in model building designated by dotted lines. Mean COVID-negative spectra (n = 38 for both edge and on crystal, at least 9 spectra per volunteer) are shown in black and COVID-positive spectra (n = 33 for edge, 31 for on crystal, at least 9 spectra per volunteer) are shown in red. Bottom frame shows the standardized Raman spectra, where each individual feature has 0 mean and unit variance. (B, E) Receiver operating curve (ROC) for these models with sensitivity and specificity. (C, F) List of features used in model building and their assignments as determined using compounds in model saliva and from literature.

Key features used in the edge model for females and males together (Figure 7C) included peaks corresponding to mucin (1146 and 1248-1249 cm^-1^), carotenoids (1146, 1170, 1580 cm^-1^) as well as multiple bands that corresponded to the amide I and III regions in proteins and nucleic acids. In the “on crystal” region, the top features used to produce the classification model included peaks associated with lipids (1123-1124, 1265-1267, 1447-1449, 1661-1663 cm^-1^), proteins (all peaks except 1643 cm^-1^), nucleic acids and urea or uric acid (1643 cm^-1^). One of the key peaks for the “on crystal” region is the peak at 1339-1340 cm^-1^ corresponding to nucleic acids. This is also a peak that is much lower and less distinct in “edge” regions, suggesting that the regions yield complementary information. Furthermore, only the peaks at 1447-1449 and 1661-1663 cm^-1^ were shared features between “edge” and “on crystal” predictive models for both sexes together (Figure 7C&F).

### 3.7 Assessing effects of potential confounding factors

Building separate models for females and males resulted in higher predictive accuracy than models including both sexes, so we investigated whether we could discriminate between samples based on sex at birth in COVID-negative samples only. The predictive model produced an ROC with an AUC of 0.70 on edge (n = 20 males, n = 18 females, MILDM) yielding 69% sensitivity, 70% specificity and 0.81 on crystal yielding 82% sensitivity, 69% specificity (MILES) (Supplementary Figure S7D-F).

We had hypothesized that Raman spectroscopy could be used to discriminate between samples from volunteers with respiratory symptoms compared to those without respiratory symptoms, perhaps as the mucus content of saliva could differ between the two. However, we found that we could not build models with high predictive accuracy to discriminate between samples based on whether reported symptoms were considered “respiratory symptoms” or not (Supplementary Figure S7A-C). We used samples from all volunteers and grouped “non-respiratory” symptomatics and asymptomatic samples together into a “non-respiratory” group and compared the corresponding spectra to those from “respiratory” symptometics (*n* = 44). Using spectra taken from all volunteers, the AUC was 0.57 in models built using “edge” spectra (*n* = 44 respiratory, *n* = 23 non-respiratory) and the AUC was 0.56 using discriminative MILES on “on crystal” spectra (*n* = 43 respiratory, n = 23 non-respiratory).

## 4 Discussion

We were able to discriminate between COVID-positive and COVID-negative saliva samples in a primer-free, label-free way using Raman spectroscopy and machine learning. We achieved a sensitivity of 79% and specificity of 75% in detecting COVID infection in non-hospitalized males with a range of symptoms, ages, medical conditions and viral load. We achieved a sensitivity of 84% and specificity of 65% in detecting COVID in non-hospitalized females with similar clinical characteristics. We found that for males, the best classification model was built using the “edge” region using features associated with carbohydrates, carotenoids, proteins and nucleic acids whilst for females, the best classification model was built using the “on crystal” region using features associated with lipids, proteins and nucleic acids. The “off crystal” region - the central region of the saliva droplet outside of the crystals - was not useful, suggesting that compounds associated with COVID-19 were perhaps less concentrated in this region than within the crystals themselves or the edge region.

We have found that the Raman spectrum of the “edge” region of dried saliva supernatant differs from that of the central region, with the central region containing peaks due to salts and proteins, whilst the edge region is primarily composed of peaks due to proteins. This is in agreement with electron microscopy studies of dried solutions of lysozyme and salt mixtures in which lysozyme was shown to accumulate at the edge of dried droplets ^48^. Although many studies claim to use Raman spectroscopy of saliva to diagnose diseases, this is the first published instance to our knowledge in which both “edge” and center regions (both “on-crystal” “off-crystal”) have been used in a single study, yielding complementary information in terms of interrogated biomolecular vibrational modes. Moreover, we could not find any other publications in which a saliva model was used to confidently assign molecular features in a Raman spectrum.

The observation that the best model for males is built using different features and spectra from different parts of the droplet compared to females suggests that COVID-19 may elicit different changes in the biomolecular profile of saliva between the sexes. Studies have shown that there are differences in the immune response of males and females to COVID, with females mounting a more robust T cell response whilst males had higher cytokine levels and are more severely impacted by COVID-19 ^49^. Furthermore, ACE2 is expressed in lower levels in liver and lung tissue of women compared to men, which could result in different impacts on metabolism ^50^.

We found that sex at birth was a confounding factor in saliva-based COVID-19 detection using label-free Raman spectroscopy as the AUC was reduced in models built from males and females together compared to those built from males and females separately. We could also discriminate between saliva supernatant samples based on sex at birth from COVID-negative females and males with an AUC of 0.81 using the “on crystal” region. This is consistent with a study by Muro et al. in which Raman spectroscopy was used to discriminate between 60 samples of whole saliva from males and females ^51^. Moreover, the biomolecular composition of saliva has been shown to be different in females and males by nuclear magnetic resonance (NMR) spectroscopy ^52^. Levels of glycine, lactic acid and acetate were all higher in saliva taken from males.

It seems we cannot discriminate between samples from volunteers with respiratory symptoms compared to volunteers without respiratory symptoms with high sensitivity or specificity using Raman spectroscopy. This may be because the majority of mucus was pelleted along with food debris during the saliva centrifugation process or because mucus content does not differ significantly in saliva supernatant between “respiratory” and “non-respiratory” samples. However, as this study occurred during December 2020-February 2021, some volunteers may have exaggerated their symptoms during reporting to obtain a PCR test for non-essential reasons e.g. travel or meeting family members during the holiday season. Unreliable reporting of symptoms is a key factor to be taken into account during infectious disease research and testing. With larger sample sizes, we may be able to determine whether the sensitivity and specificity of our test is affected by whether the volunteer has respiratory symptoms, non-respiratory symptoms or no symptoms.

## 5 Conclusions

In conclusion, we showed that Raman spectroscopy can be used to detect biomolecular changes between COVID-positive and COVID-negative saliva supernatant and that accounting for the sex of the saliva donor can increase the accuracy of predictive models. However, limitations of our Raman microspectroscopy approach include the fact that it was only possible to sample less than 1% of the full sample area and imaging times were too long for rapid COVID screening (2-3 hours per drop). It is likely that in this study, we may not have captured the full molecular profile of every saliva drop. Therefore, we have developed in parallel a rapid single-point Raman spectroscopy platform similar to the probe we have already developed and commercialized for detecting brain cancer ^20,53,54^. This can image a whole droplet within a few seconds and is portable, affordable and suitable for high throughput screening. The platform could also be used for detection of other infectious diseases or COVID variants simply by retraining the machine learning algorithms on new samples.

Moreover, saliva is a complex medium and multiple factors can affect the Raman spectrum. However, even whilst taking into account sex at birth, the maximum AUC in our study was 0.80. This suggests that there are further confounding factors. We are currently expanding our analysis to image the remaining 475 COVID-negative samples to take such factors into account using the single point platform. We will be thoroughly assessing the effects of more confounding factors such as age, diet, smoking and comorbidities on the salivary Raman fingerprint and evaluating whether these variables can impact the accuracy of detecting COVID-19. We will also look further into factors that could affect hormone levels such as pregnancy, medical conditions, prescription medications and surgeries, and assess their impact on the Raman spectrum of saliva.

## Data Availability

Data is available upon request from the authors.

## 6 Notes and references

### 6.1 Author Contributions

Conceptualization: KE, FD1, DT, FL

Methodology: KE, FD1, MM, TN, DT, FL

Ethics: MM, KE

Maintenance of microscope: GS

Sample collection: KE, MM, EZ, TT, GB

Sample processing: KE, MM, EZ, TT

Model saliva preparation: MKD

Data acquisition: KE, FD1, TN, MM, EZ, TT,

Data acquisition from model saliva and components: JL, AF, KE, FD1, TN

Visualization: FD2, AP

Data analysis: FD2, AP, FL Data interpretation: KE, FL

Supervision: FL, DT

Writing and editing: KE, EZ, TT, AS, FD2, NK, FL, CQ

KE: Katherine Ember
FD1: François Daoust
MM: Myriam Mahfoud
FD2: Frédérick Dallaire
EZ: Esmat Zamani
TT: Trang Tran
AP: Arthur Plante
MKD: Mame-Kany Diop
TN: Tien Nguyen
AS: Amélie St-Georges-Robillard
NK: Nassim Ksantini
JL: Julie Lanthier
AF: Antoine Filiatrault
GS: Guillaume Sheehy
CQ: Caroline Quach,
DT: Dominique Trudel
FL: Frédéric Leblond

## 6.3 Acknowledgements

We would like to acknowledge Julie Dionne and Axel Bergman from the TransMedTech team. We would like to thank Laurence Knafo for biohazard training and access to containment level 2 facilities. We would like to thank the following personnel for assistance with questionnaire data entry: Gabriel Beaudoin, Jérémie Kerouac, Thomas Regouffre, Jean-Francois Martin. We would also like to thank all staff at the Pointe St-Charles testing center, especially Jacynthe, David and Samia.

