## Supplemental Material for "Saliva-based detection of COVID-19 infection in a real-world setting using reagent-free Raman spectroscopy and machine learning"

† First author
‡Equally contributing authors

**This PDF file includes:**

Figs. S1 to S8
Tables S1 to S4
References (1 to 5)

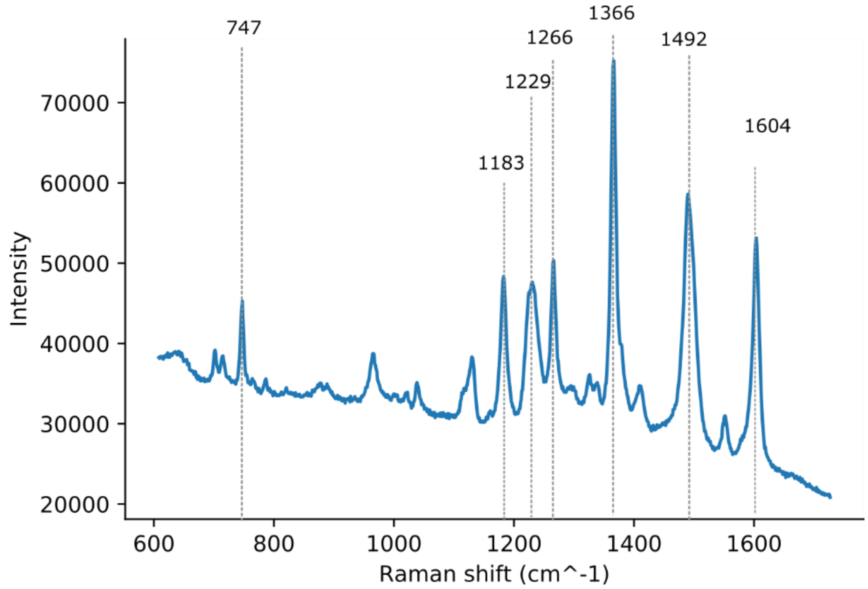


**Fig. S1.**

A Raman spectrum from lipstick contamination in a saliva supernatant sample from a COVID-19 negative volunteer. Although the saliva donor was wearing a red lipstick from Max factor, the Raman spectrum is remarkably similar to that of red Bourjois 15 (pure and not in saliva) which has peaks at or one wavenumber away from 747, 1183, 1229, 1266, 1366, 1492 and 1604 $cm^{-1}$.

**Table S1.**
Available viral load data of 18 COVID positive samples (sample numbers are arbitrary)

| Sample | Ct Target 1 | Ct Target 2 |
|---|---|---|
| 1 | 18.2 | 18.2 |
| 2 | 30.6 | 30.6 |
| 3 | 19.2 | 19.4 |
| 4 | 18 | 18 |
| 5 | 27.1 | 27.1 |
| 6 | 17.9 | 17.6 |
| 7 | 33.5 | 35.6 |
| 8 | 20.2 | 20.2 |
| 9 | 21.7 | 22.4 |
| 10 | 20.2 | 20.3 |
| 11 | 24.1 | 23.8 |
| 12 | 15.5 | 15.5 |
| 13 | 32.7 | 34 |
| 14 | 0 | 35.3 |
| 15 | 33.8 | 35.4 |
| 16 | 0 | 36.3 |
| 17 | 32.6 | 34.5 |
| 18 | 20.1 | 20.5 |

**Table S2.**
Concentrations of compounds added to distilled $H_2O$ to produce model saliva.

| **Compound** | **Concentration (g/L)** |
|---|---|
| Sodium chloride | 1.594 |
| Ammonium nitrate | 0.057 |
| Potassium phosphate | 0.636 |
| Potassium chloride | 0.202 |
| Potassium citrate | 0.308 |
| Uric acid sodium salt | 0.021 |
| Urea | 0.198 |
| Lactic acid sodium salt | 0.146 |
| Glucose | 0.014 |
| Human serum albumin | 4 |
| Bovine submaxillary mucin | 1 |

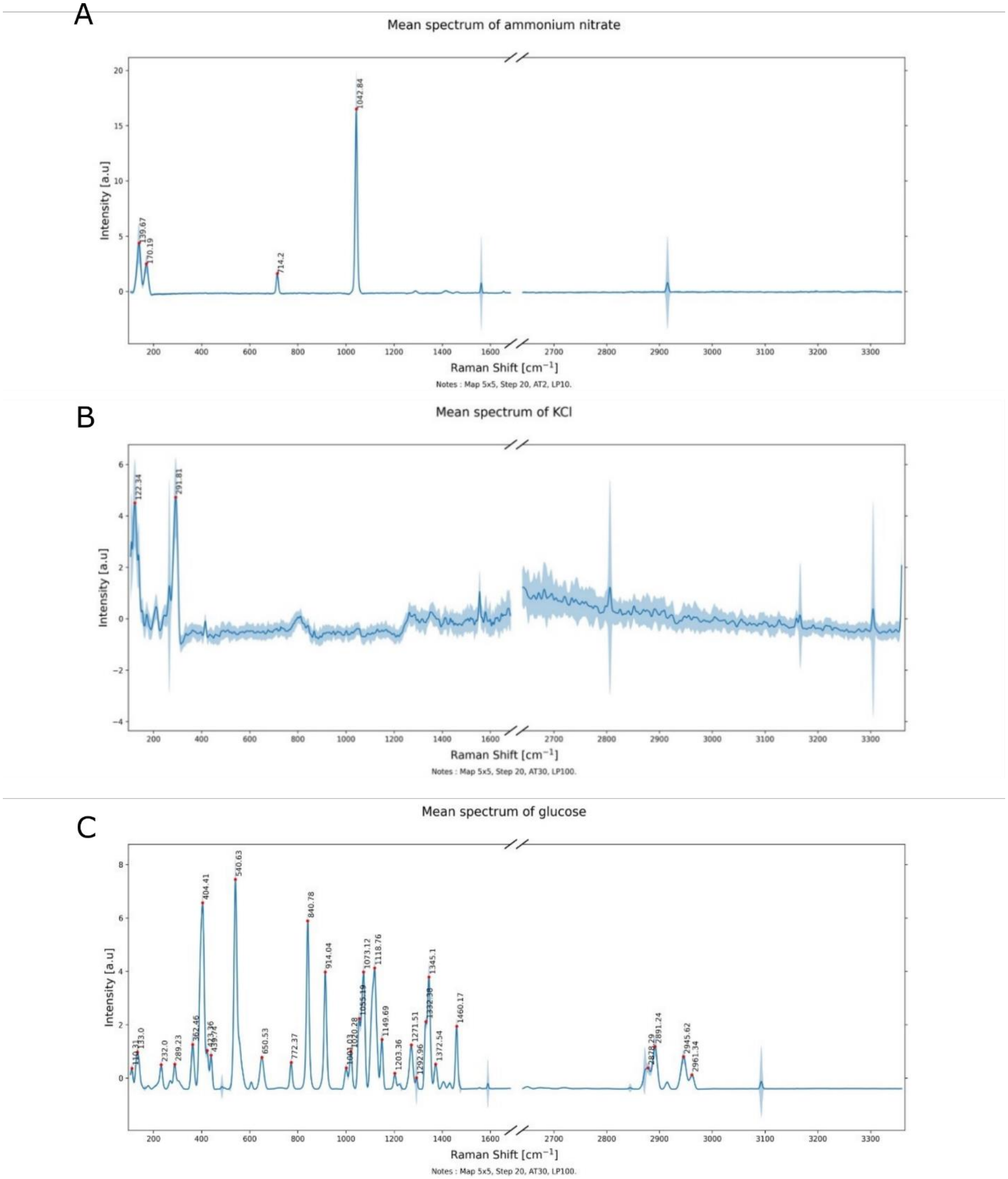


**Fig. S3.**
Raman spectra from a) ammonium nitrate, b) potassium chloride and c) glucose. Spectra were taken using 785 nm excitation with a Renishaw InVia Raman spectrometer from solid compounds placed on an aluminum slide.

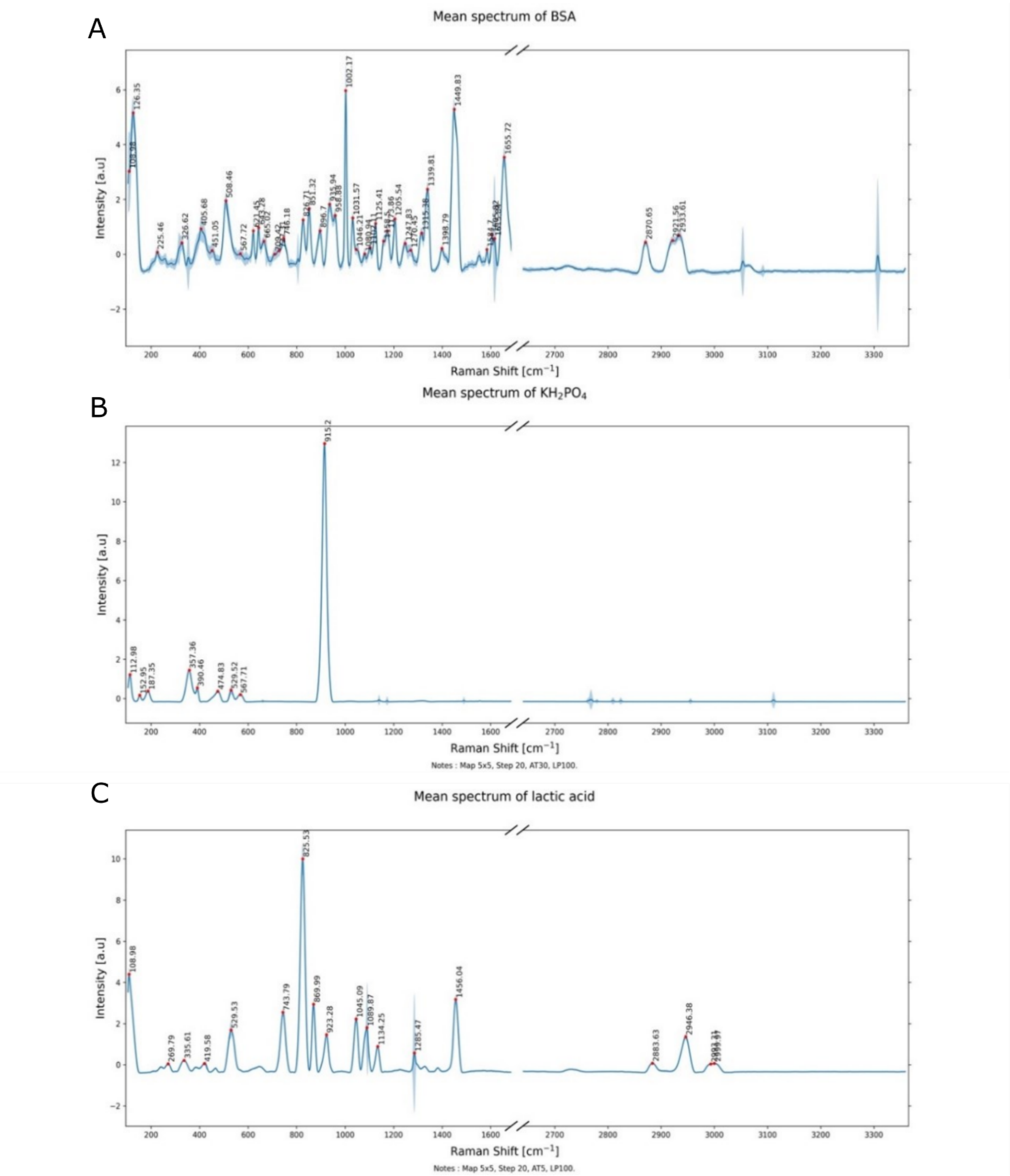


**Fig. S4.**

Raman spectra from a) bovine serum albumin, b) potassium phosphate and c) lactic acid. Spectra were taken using 785 nm excitation with a Renishaw InVia Raman spectrometer from solid compounds placed on an aluminum slide.

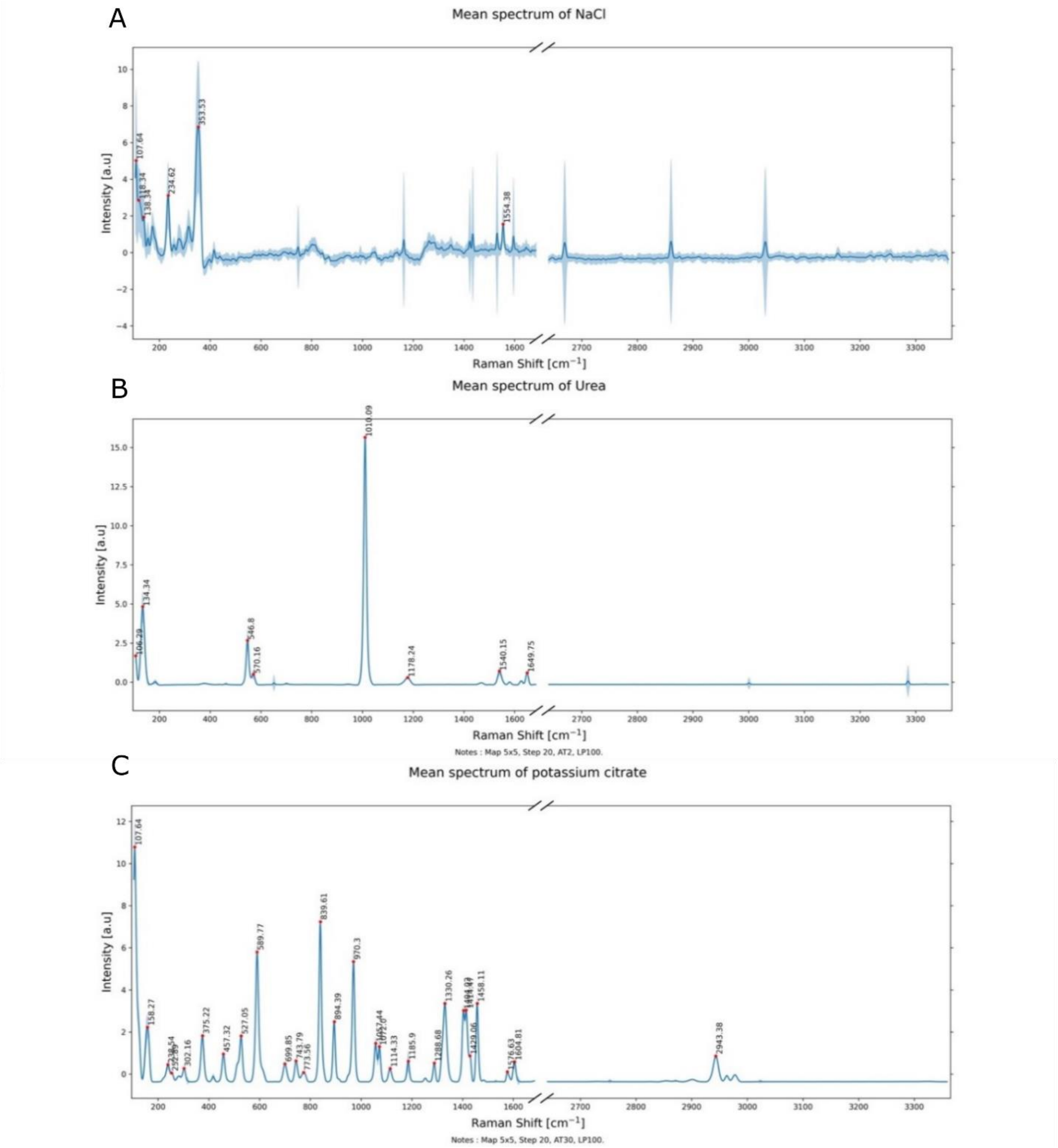


**Fig. S5.**
Raman spectra from a) sodium chloride, b) urea and c) potassium citrate. Spectra were taken using 785 nm excitation with a Renishaw InVia Raman spectrometer from solid compounds placed on an aluminum.

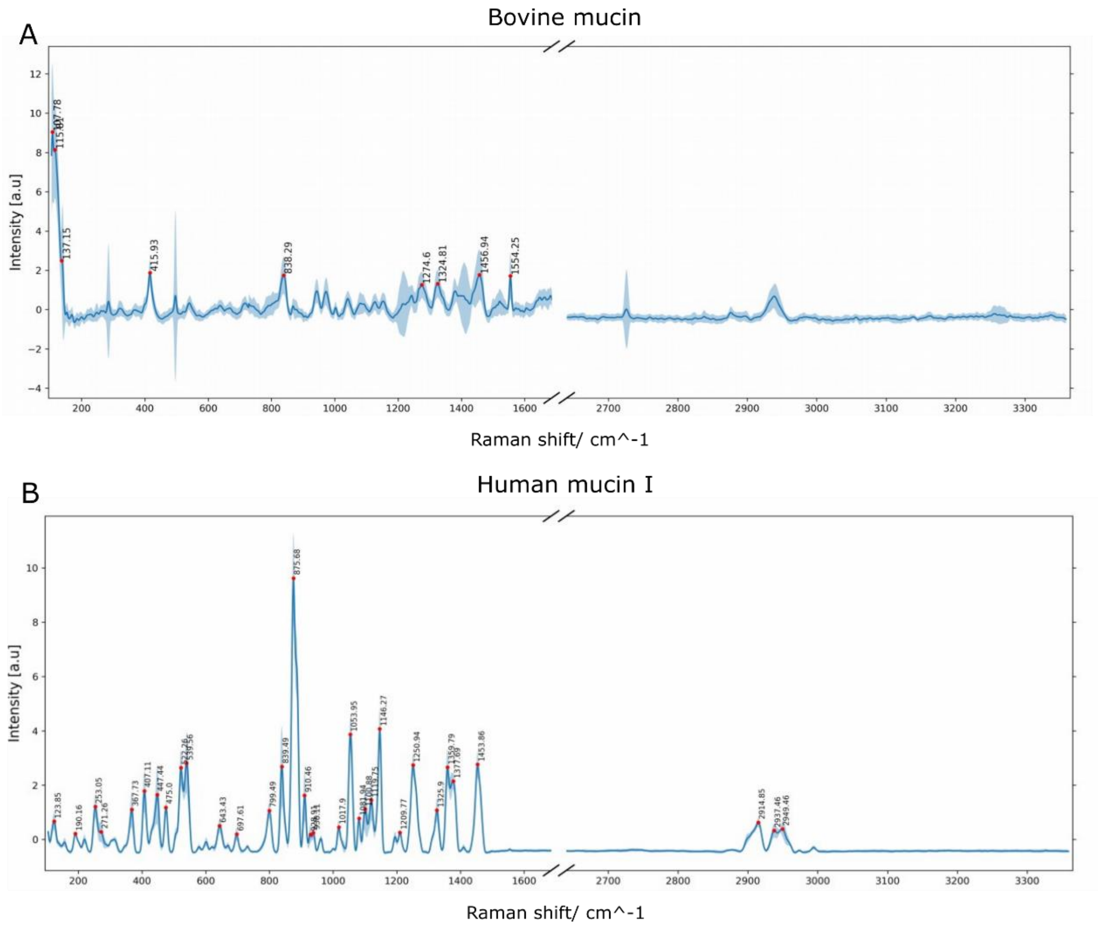


**Fig. S6.**
Raman spectra from a) bovine submaxillary mucin and b) human mucin I. Spectra were taken using 785 nm excitation with a Renishaw InVia Raman spectrometer from solid compounds placed on an aluminum slide.

**Table S3.**
Table of the Raman peaks present in Raman spectra from a dried droplets of model saliva and human saliva supernatant (shown in Figs 2 and 3 of main paper) in addition to their location (presence in edge or center of the droplets) and the assignments from model saliva constituents. "Center" refers to on crystal and off crystal regions as peaks were present in both. Additional possible contributions to bands from literature are taken from [2–5]. Raman peaks for some molecules may be undetectable as the peaks are either not present with sufficient signal to noise ratio, or because the peaks lie under the shoulders of broad adjacent peaks.

| **Raman peak / $cm^{-1}$** | **Location of peak in model saliva** | **Location of peak in human saliva supernatant** | **Biomolecular assignments** | |
|---|---|---|---|---|
| | | | **Band assignment based in peak position from consituents of model saliva** | **Additional possible contributions to bands from literature** |
| 623 | Edge | Edge, Center | Protein (phenylalanine) or uric acid | |
| 646 | Edge | Edge, Center | Protein (tyrosine, phenylalanine), glucose | |
| 718 | Undetected | Edge, center | | DNA, phospholipids |
| 731 | Undetected | Edge | | Adenine, phosphatidylserine |
| 746 | Edge | Center | Protein | DNA |
| 760 | Undetected | Edge, center | | Tryptophan, ethanolamine |
| 790 | Center | Edge, center | Glucose, uric acid | Phosphodiester |
| 805 | Undetected | Edge, center | | Uracil |
| 827 | Undetected | Edge, center | Protein | Phosphodiester in DNA |
| 835 | Edge | Undetected | | Amine |
| 853 | Edge, center | Edge, center | Glucose, potassium citrate | Tyrosine, proline, polysaccharides |
| 876 | Undetected | Edge, center | Phosphate, human mucin I | Choline, phospholipids |
| 890 | Undetected | Center | | Proteins, carbohydrates |
| 900 | Edge | Undetected | | Carbohydrates |

| | | | | |
|---|---|---|---|---|
| 924 | Undetected | Edge, center | Phosphate, glucose and protein (proline), lactic acid | |
| 935 | Center | Edge | Protein (proline, valine) | Glycogen |
| 942 | Edge | Undetected | | Carbohydrates |
| 957 | Edge | Center | Protein | Hydroxyapetite, carotenoid, cholesterol |
| 1003 | Center and edge | Center and edge | Phenylalanine/protein | NADH |
| 1031 | Center and edge | Edge | Protein (phenylalanine) | Phospholipids |
| 1045 | Edge (weak), center | Edge (weak), center (strong) | Nitrate and protein (phenylalanine), uric acid, lactic acid, human mucin I | Phosphate, carbohydrate |
| 1082 | Edge, center (weak) | Center (weak) | Protein, glucose | Carbohydrates, nucleic acids, phospholipids, ATP |
| 1101 | Edge | Edge | Protein | C-N, lipids |
| 1112 | Undetected | Edge | | Carbohydrates, carotenoids |
| 1125 | Edge, center (weak) | Edge, center | Protein | Lipid, RNA (ribose), carbohydrate, blood, porphyrin |
| 1146 | Edge | Edge, center | Human mucin I | Carbohydrates, carotenoids |
| 1156 | Edge | Edge, center | Protein | Carotenoids |
| 1173 | Edge, center (weak) | Edge | Protein (tyrosine), urea | Carotenoids |
| 1203 | Center (weak) | Center (weak) | | Nucleic acids, amide III |
| 1206 | Edge | Edge | Protein (amide III) | Nucleic acids, IgG |
| 1250 | Undetected | Edge | Protein (amide III), human mucin I | Asymmetric phosphate, DNA/ RNA (guanine, cytosine) |
| 1266 | Undetected | Edge, center | Protein (amide III) | Nucleic acids, fatty acids |
| 1319 | Edge | Edge, center | Protein (amide III) | Nucleic acids (guanine) |

| | | | | |
|---|---|---|---|---|
| 1338 | Edge and center | Edge, center | Protein (amide III) | Nucleic acids |
| 1401 | Center | Edge | | Methyl groups, aspartate, glutamate |
| 1417 | Center | Edge (weaker), center (stronger) | | Aspartate, glutamate |
| 1449 | Edge (strong), center | Edge and center | Protein (amide I), lactic acid | Lipids, red blood cells, aromatic carbonds |
| 1512 | Undetected | Edge, center | | DNA, cytosine |
| 1519 | Undetected | Edge | | Carotenoid, porphyrin |
| 1553 | Center (strong relative to edge), edge (v weak) | Edge, center | Protein (tryptophan, amide II), sodium chloride | Mucin, porphyrin |
| 1574 | | Edge | | DNA |
| 1584 | Edge | Edge | Citrate | Phenylalanine, ATP, carotenoids, DNA/RNA |
| 1600 | Center | Undetected | | Amide I, phenylalanine |
| 1605 | Edge | Edge | Protein (amide I) | DNA |
| 1616 | Edge | Edge | Protein (tyrosine, tryptophan) | Porphyrin |
| 1655 | Edge (strong), center (v weak) | Undetected | Protein (amide I), urea, uric acid | Lipid |
| 1665 | Center | Edge | Protein (amide I) | Unsaturated fatty acids, DNA |

**Table S4.**

Area under curve (AUC) values for receiver operating characteristic (ROC) curves produced for predictive models generated using both MILES and MILDM in the study. "Crop" refers to spectra that have had the region with high variance before 1100 $cm^{-1}$ removed.

| Spectra type | Regions | Sex | Classification | COVID status | Nb patients | Nb spectra | AUC MILES | AUC MILDM |
|---|---|---|---|---|---|---|---|---|
| All | Edge | Both | Covid status | All | 71 | 702 | 0.676 | 0.671 |
| Crop | On crystal | Both | Covid status | All | 69 | 687 | 0.69 | 0.628 |
| Crop | Off crystal | Both | Covid status | All | 69 | 668 | 0.592 | 0.571 |
| Crop | Edge | Both | Covid status | All | 71 | 702 | 0.709 | 0.757 |
| Crop | On crystal | Male | Covid status | All | 35 | 347 | 0.589 | 0.722 |
| Crop | Edge | Male | Covid status | All | 35 | 342 | 0.735 | 0.800 |
| Crop | On crystal | Female | Covid status | All | 34 | 300 | 0.796 | 0.789 |
| Crop | Edge | Female | Covid status | All | 36 | 320 | 0.673 | 0.651 |
| Crop | On crystal | Both | Symptoms (respiratory vs. non-respiratory) | All | 69 | 687 | 0.551 | 0.587 |
| Crop | Edge | Both | Symptoms (respiratory vs. non-respiratory) | All | 71 | 702 | 0.559 | 0.612 |
| Crop | On crystal | Both | Sex | Negative | 37 | 687 | 0.805 | 0.669 |
| Crop | Edge | Both | Sex | Negative | 38 | 702 | 0.694 | 0.704 |

Edge

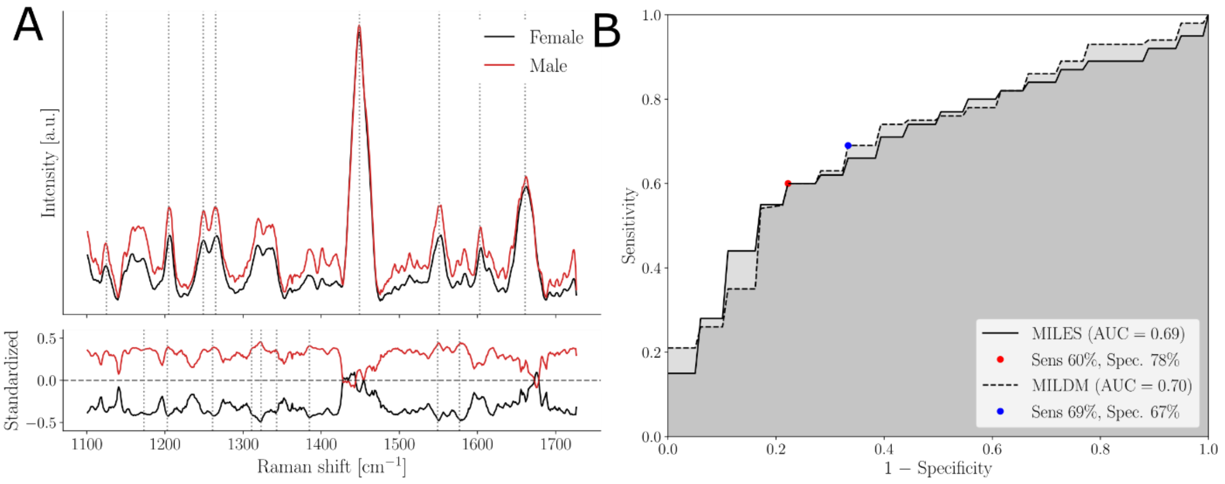


C

| Peak center (cm$^{-1}$) | Model saliva correspondence | Biomolecular assignement |
|---|---|---|
| 1123–1124 | Protein | RNA (ribose), lipids, carbohydrates, porphyrin, blood |
| 1203–1206 | Protein (amide III) | DNA/RNA, IgG |
| 1248–1249 | Protein (amide III), mucin | Asymmetric phosphate, DNA/RNA(guanine, cytosine) |
| 1265–1267 | Protein (amide III) | DNA/RNA, fatty acids |
| 1447–1449 | Protein (amide I), lactic acid | Lipids, RBCs |
| 1552–1553 | Protein(tryptophan), sodium chloride | Mucin, porphyrin |
| 1603–1605 | Protein (amide I) | DNA |
| 1661–1663 | Protein (amide I) | Unsaturated fatty acids, DNA |

On crystal

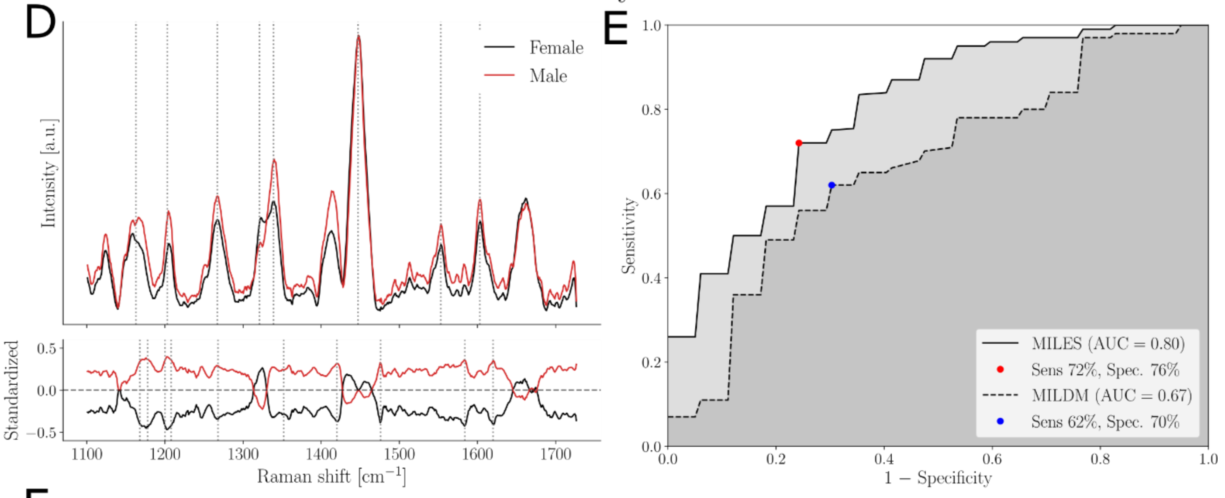


F

| Peak center (cm$^{-1}$) | Model saliva correspondence | Biomolecular assignement |
|---|---|---|
| 1163–1164 | Protein | Mucin, quinoid, lipids |
| 1203–1206 | Protein (amide III) | DNA/RNA, IgG |
| 1265–1267 | Protein (amide III) | DNA/RNA, fatty acids |
| 1321–1323 | Protein (amide III) | DNA/RNA (guanine) |
| 1339–1340 | Protein | DNA/RNA |
| 1447–1449 | Protein (amide I), lactic acid | Lipids, RBCs |
| 1552–1553 | Protein(tryptophan), sodium chloride | Mucin, porphyrin |
| 1603–1605 | Protein (amide I) | DNA |

**Fig. S7.**

Machine learning model discriminating between female and male saliva supernatant from COVID-negative volunteers using (A-C) “edge” and (D-F) “on crystal” Raman spectra from dried droplets: (A, D) Upper frame shows SNV-normalized, baseline corrected Raman spectra from all volunteers. Variance is shown by pale lines (variance of mean spectrum from each individual) and main features used in model building designated by dotted lines. Mean female spectra (n = 18, at least 9 spectra per volunteer) are shown in black and mean meale spectra (n = 20, at least 9 spectra per volunteer) are shown in red. Bottom frame shows the standardized Raman spectra, where each individual feature has 0 mean and unit variance. (B, E) Receiver operating curve (ROC) for these models with sensitivity and specificity. (C, F) List of features used in model building and their assignments as determined using compounds in model saliva and from literature.

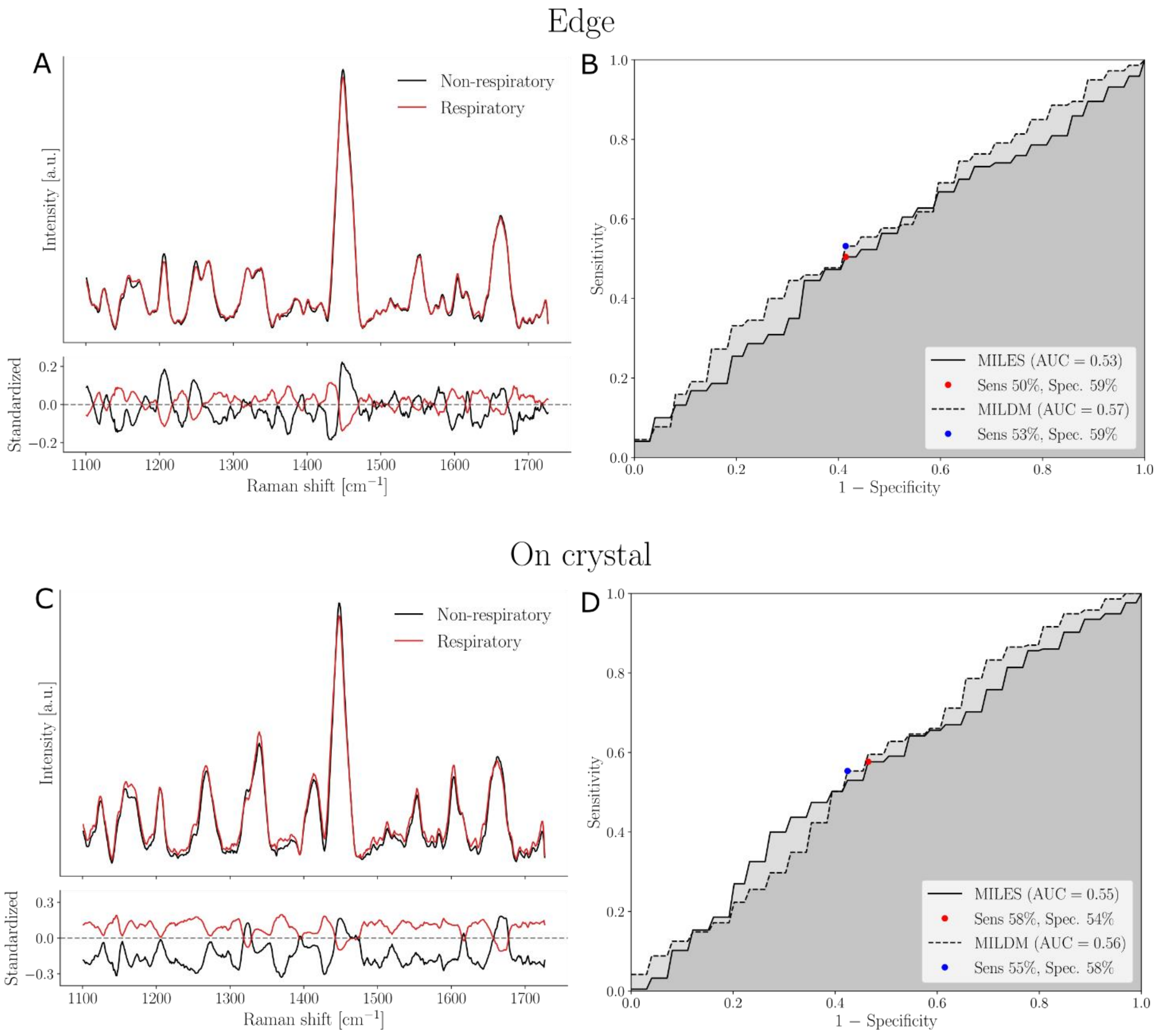


**Fig. S8.**

Machine learning model discriminating between respiratory and non-respiratory saliva supernatant from volunteers using (A-B) "edge" and (C-D) "on crystal" Raman spectra from dried droplets: (A, B) Upper frame shows SNV-normalized, baseline corrected Raman spectra from all volunteers. Variance is shown by pale lines (variance of mean spectrum from each individual) and main features used in model building designated by dotted lines. Mean non-respiratory spectra (*n* = 23, at least 9 spectra per volunteer) are shown in black and respiratory spectra (*n* = 44 for edge, 43 for on crystal at least 9 spectra per volunteer) are shown in red. Bottom frame shows the standardized Raman spectra, where each individual feature has 0 mean and unit variance. (C, D) Receiver operating curve (ROC) for these models with sensitivity and specificity.
